# Sub-national heterogeneity in the time-varying reproduction number during the 2026 Bundibugyo virus disease outbreak in the Democratic Republic of the Congo: a hierarchical Bayesian analysis

**DOI:** 10.64898/2026.08.19.26360792

**Authors:** Johan G. L. Verheyden, Celestin Nzanzu Mudogo, Wolfgang Jacquet

**Affiliations:** Aries Consult Kenya, Nairobi, Kenya; Faculté de Médecine, Université de Kinshasa, Kinshasa, Democratic Republic of the Congo; Departement Clinical Sciences, Faculty of Medicine and Pharma, Vrije Universiteit Brussel

**Keywords:** Bundibugyo virus disease, Ebola, reproduction number, Bayesian hierarchical model, renewal equation, Democratic Republic of the Congo, disease surveillance

## Abstract

National-level estimates of the time-varying reproduction number (Rt) for the 2026 Bundibugyo virus disease (BDBV) outbreak in the Democratic Republic of the Congo (DRC) declined from a value close to the epidemic threshold in early August 2026 to a modestly sub-threshold value by late August, consistent with an independent case-count series compiled separately from the same underlying situation reports. A single national Rt, however, can obscure divergent sub-national epidemic trajectories, particularly across an outbreak that had by late August reached six provinces, ranging from a declining original epicentre to recently-seeded fronts. We estimated Rt at national, provincial, and — where case volume allowed — health-zone level, using both a standard sliding-window (Cori) estimator and a hierarchical Bayesian renewal model with partial pooling across spatial units, fitted by Hamiltonian Monte Carlo (No-U-Turn Sampler), with a province- or zone-specific (rather than shared) dispersion parameter retained on the basis of leave-one-out cross-validation. As of the week of 20–26 August 2026, Ituri — the outbreak’s original epicentre — had a hierarchical median Rt of 0.79 (95% credible interval [CrI] 0.61–1.04), Nord-Kivu remained above threshold (1.10, [0.89–1.39]), and Haut-Uele, now individually resolved at health-zone level for the first time, sat near threshold (1.01, [0.69–1.38]). Health-zone disaggregation, feasible in Ituri, Nord-Kivu, and now Haut-Uele given case volume, showed the provincial picture itself masked further heterogeneity, and that this heterogeneity is not static: Mongbwalu, the zone in which the outbreak began, remained in clear decline (Rt 0.37, [0.19–0.74]), while Rwampara — elevated three weeks earlier — had reversed to a clearly sub-threshold trajectory (0.53, [0.35–0.80]); conversely Butembo, previously in decline, had reversed to an elevated trajectory (1.36, [0.85–2.07]) comparable to the persistently-elevated Katwa (1.09, [0.82–1.41]). An initial disagreement between the sliding-window and hierarchical provincial estimates was traced to a data-reconstruction artefact (forward-filling, rather than interpolating, multi-day gaps in health-zone reporting) rather than a genuine methods disagreement; a targeted validation experiment, injecting an equivalent reporting gap into an independent historical outbreak’s case series (Beni health zone, 2018–2020 North Kivu/Ituri epidemic), confirmed that linear interpolation reduces reconstruction error roughly five-fold relative to forward-fill under the same failure mode, indicating the correction generalises beyond this specific outbreak. The hierarchical model achieved 96.1% pooled 95% posterior-predictive interval coverage, and the provincial ranking was unchanged across a generation-interval sensitivity grid (Spearman ρ = 1.0). Aggregation masks meaningful heterogeneity in transmission intensity at every spatial resolution examined, and that heterogeneity itself shifts over periods as short as two to three weeks; response prioritisation based on a single national, provincial, or even a single dated health-zone snapshot risks directing attention away from where transmission is currently supercritical.

**Author Summary:** During an Ebola outbreak, public health teams track a number called the reproduction number, or Rt — roughly, how many new people each infected person goes on to infect. When Rt is above 1, an outbreak is growing; below 1, it is shrinking. For the 2026 Bundibugyo virus disease outbreak in the Democratic Republic of the Congo, the national Rt has moved from close to 1 in early August 2026 to modestly below 1 by late August. We show that this single national number hides very different, and rapidly changing, realities in different places. The province where the outbreak started, Ituri, has an estimated Rt below 1 overall — but even within Ituri, specific towns move independently: one area that was still growing three weeks ago has since turned around, while the original outbreak site continues to decline. In Nord-Kivu province, one town that had been declining has since reversed and is now growing again, while a neighbouring town remains elevated throughout. We used a statistical technique called hierarchical Bayesian modelling, which allows data-poor areas to “borrow strength” from data-rich ones rather than producing wild, uninterpretable estimates, and which we validate three separate ways, including by deliberately reproducing a real data problem in a completely different, older outbreak’s data to check that our fix for it generalises. We also found and corrected a data-processing error that had made a simpler method appear to disagree with our more careful one — a reminder that methodological rigour includes checking the data pipeline, not just the statistical model. Our results suggest that outbreak response resources should be targeted at the specific health zones still driving transmission, and reassessed frequently, since a national, provincial, or even a single dated health-zone reading can be overtaken by events within a few weeks.

## 1. Introduction

The Democratic Republic of the Congo (DRC) has, since 15 May 2026, been responding to its seventeenth recorded Ebola epidemic and only the second caused by Bundibugyo virus (BDBV) [1]. By late August 2026 the outbreak had produced nearly 6,000 confirmed cases across six provinces, with a case-fatality ratio exceeding the historical range reported for prior BDBV outbreaks [2]. Real-time estimation of the time-varying reproduction number, Rt — the average number of secondary infections produced by an infectious case at time t, given the intervention and immunity conditions prevailing at that time [3] — is a standard input to outbreak response planning, indicating whether transmission is currently supercritical (Rt > 1) or in decline (Rt < 1).

Existing Rt estimates for this outbreak are national in scope. A live joint Bayesian renewal-process model [4] fits multiple national surveillance streams — confirmed and suspected cases, deaths, laboratory throughput, treatment-centre occupancy, and cross-border exports — with modelled reporting delays and ascertainment, and has tracked Rt since May 2026. National scenario projections from a separate transmission model have also been published [5]. Neither approach, nor any published analysis we are aware of, has examined whether a single national Rt adequately characterises transmission across an outbreak spanning six provinces and, within the most-affected province, dozens of health zones — the sub-provincial administrative and surveillance unit around which DRC’s health system is organised, broadly analogous to a health district — with widely differing, and independently evolving, epidemic histories.

This matters for three related reasons. First, statistically: if provinces or health zones have genuinely different Rt trajectories, a national estimate is a case-weighted average that can sit near the epidemic threshold even while some areas are clearly declining and others are clearly still growing. Second, operationally: response prioritisation decisions (where to reinforce contact-tracing capacity, treatment-centre bed capacity, or community engagement) are made at the province and health-zone level, not the national level, and are better served by an estimate at the resolution the decision is actually made. Third, temporally: as we show directly by extending our own analysis window by two and a half weeks over the course of this study, zone-level transmission intensity can reverse direction within that span — a single dated snapshot, however granular, is therefore itself a limited guide to current risk.

We address this by estimating Rt hierarchically at national, provincial, and (where supported by case volume) health-zone resolution, comparing a hierarchical Bayesian renewal model against a standard sliding-window estimator throughout, reporting a data-reconstruction artefact we identified and corrected in the course of this comparison as a methodological finding in its own right, and testing whether the correction for that artefact generalises to a different outbreak’s data.

### 1.1 Relationship to companion analyses

This paper draws on the same INRB-UMIE surveillance infrastructure [6] as a wider portfolio of companion analyses from our group, all now publicly posted [7–14]. A spatial hazard model of which health zone will next report a first confirmed case [7] addresses a distinct question — invasion into currently-unaffected zones, evaluated by rolling-origin rank-based hit rate — from this paper’s focus on transmission intensity within already-affected units, evaluated by a renewal-equation likelihood; we confirmed no methodological or content overlap by direct comparison. A national short-term case-forecasting analysis [8] forecasts future case and death counts by rolling-origin evaluation of a recalibrated Bayesian negative-binomial model, a phenomenological growth-curve benchmark, and a data-driven ensemble, assessed by predictive-interval coverage and weighted interval score rather than by transmission-intensity estimation; we confirmed by direct comparison that it does not estimate Rt at any resolution and does not overlap this paper.

An analysis of health-zone case-fatality ratio measurement [9] fits beta-binomial partial-pooling models to national and health-zone confirmed cases and deaths through 20 July 2026, examining reported crude case-fatality heterogeneity, its association with epidemic maturity and mapped clinical-facility context, and case-death reporting synchronisation. It explicitly does not estimate a biological fatality risk, and does not estimate Rt, growth rate, or any transmission-intensity quantity at any resolution; its unit of analysis (health-zone deaths-to-cases ratio) and its identifiability argument (a reported ratio conflates ascertainment, reporting delay, and outcome maturity with the underlying biological quantity) are conceptually adjacent to this paper’s own report-date-vs-onset-date caveat and our new reporting-capacity caveat (both Section 4.4) but address mortality, not transmission; we confirmed no material overlap by direct reading.

Two further quantitative companion analyses address related but distinct questions. A growth-rate and back-calculation analysis [10] estimates a single exponential growth rate and doubling time over the outbreak’s early, pre-discontinuity phase (14–27 May 2026) and back-calculates transmission onset to 19 April 2026, entirely at national resolution; it deliberately does not commit to a generation-interval assumption, and so reports growth rate and doubling time rather than a reproduction number, and does not construct a time-varying Rt trajectory at any resolution. This differs from the present paper in estimand, method, and spatial resolution; we confirmed no material overlap by direct comparison of both manuscripts. An outbreak-origin analysis [11] uses approximate Bayesian computation to compare four hypotheses for whether a retrospectively-reported January/February cluster in Mongbwalu was ancestral to the May epidemic. Its simulator parameterises a weekly reproduction number, R(t), as an internal generative-model input across four early-corridor health zones and a January–June window, but R(t) is not itself a reported estimate; its temporal scope (pre-declaration origin reconstruction) and spatial scope (four zones, one corridor, one early window) do not overlap the present paper’s full-outbreak, six-province, continuously-updated analysis. We note for completeness that three of the four zones in that paper’s early corridor (Mongbwalu, Bunia, Rwampara) also appear in our Ituri health-zone results (Table 2), addressing a different question about them (current transmission intensity, not origin) over different, later windows.

A sixth companion paper [12] develops a general estimand-first framework for judging, at a given surveillance vintage, which analytical products an outbreak dataset can responsibly support — distinguishing surveillance maturity from epidemic-process informativeness and defining six non-hierarchical inferential-support classes. Its own empirical demonstrations use the 2018–2020 North Kivu/Ituri Ebola outbreak (the same historical outbreak we draw on independently in Section 2.9 for a different purpose), an independent Sierra Leone forecasting dataset, and a synthetic mortality-identifiability experiment; the 2026 BDBV outbreak is explicitly described there as contemporary motivation rather than the principal validation dataset, and it does not itself estimate Rt for the present outbreak. We draw on it directly in Section 4.2: our own data-reconstruction artefact is a worked instance of its finding that surveillance recovery can be an analytically unstable transition.

Finally, two qualitative companion analyses — a desk-based synthesis of burial, mourning, and the social organisation of care [13] and a parallel synthesis of trust, rumour, and response legitimacy [14] during this outbreak — sit in the same portfolio but address social-science questions with no quantitative or methodological overlap with a renewal-equation transmission analysis; we note them for portfolio completeness.

## 2. Materials and Methods

### 2.1 Data sources

Confirmed-case counts through 11 August 2026 were drawn from two INRB-UMIE-maintained data products, both ultimately sourced from Institut National de Santé Publique (INSP) situation reports (“SitReps”): a vintage-by-vintage national transcription (epiforecasts/BVDOutbreakSize) and a health-zone-resolution daily contract file (INRB-UMIE/BDBV2026-Data), canonicalised to standard zone names via the repository’s own alias table and mapped to province via its shapefile-derived zone/province lookup. The INRB-UMIE/BDBV2026-Data repository is openly available with acknowledgement, citable by its Zenodo DOI [15] and the accompanying publication [6].

For the period 11–26 August 2026, health-zone case tables were extracted directly by us from the underlying INSP SitRep PDFs (SitReps 089, 093, 094, 096, 097, 099, 101, 102, and 104), since the processed health-zone product had not yet incorporated this window at the time of analysis. Extraction used layout-preserving text parsing with a purpose-built table parser, accommodating two distinct SitRep table formats encountered across this window. Every extracted province-level total was cross-validated against the same SitRep’s own independently-printed province-summary table; zone-level sums matched printed province totals exactly for four of six provinces at every date checked, with a single, immaterial one-case discrepancy in Tshopo (a province not part of the health-zone extension) at most dates. SitRep 094 (17 August) used a substantially different table template that could not be parsed with matched reliability and was excluded from the health-zone series; the resulting single-day gap is handled identically to other reporting gaps (Section 2.8). At the point of first overlap between our own extraction and the pre-existing processed product (11 August), all 47 zones present in both sources agreed exactly. Our independently-reconstructed national daily case count also agreed with a separately-compiled, more recently updated release of the national comparator series (epiforecasts/BVDOutbreakSize, updated independently of our own extraction) to within a handful of cases across eight consecutive dates in the extension window, which we take as further corroboration of the extraction pipeline. Six zone names required manual reconciliation between orthographic variants used in the original processed product and in raw SitRep text (e.g. “Nia Nia” vs. “Nia-Nia”, “Gety” vs. “Gethy”), resolved against the canonical spelling in the shapefile-derived zone list; this reconciliation is documented in the reproducibility archive (Data and Code Availability).

The final analysis uses data through 26 August 2026 throughout (national, provincial, and health-zone level alike), a single consistent cutoff, superseding the two-cutoff approach necessitated in an earlier version of this analysis by the health-zone product’s coverage lag.

### 2.2 Generation interval

No BDBV-specific generation-interval estimate exists. We used the Ebola serial-interval estimate of the WHO Ebola Response Team [16] — mean 15.3 days, SD 9.3 days — as a generation-time proxy, discretised as a Gamma distribution (shape 2.71, scale 5.65) truncated to whole-day lags with the zero-lag mass dropped and the remainder renormalised. This is the same proxy and discretisation used by the national comparator model [4], chosen deliberately for comparability. Sensitivity to this choice is assessed in Section 2.6.

### 2.3 Independent (sliding-window) Rt estimate

As a comparator at every spatial resolution, we computed the sliding-window instantaneous reproduction number of Cori et al. [3] in closed form: for a window of length τ ending on day t, R ∼ Gamma(a + ΣI, b + ΣΛ) under a Gamma(a=1, b=0.2) prior, where I is daily incidence and Λ_t = Σ w I_{t−i} is the total infectiousness on day t under the generation-interval probability mass function w. We used τ = 14 days and τ = 7 days at national level and report the posterior median and 95% credible interval.

### 2.4 Hierarchical Bayesian renewal model

For the provincial and health-zone analyses we fitted a hierarchical Bayesian model with partial pooling across spatial units p (province or, in the zone-level extension, health zone). Rt was modelled as a unit-specific weekly random walk on the log scale:

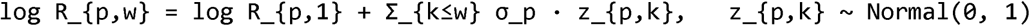

with the random-walk step size itself partially pooled across units,

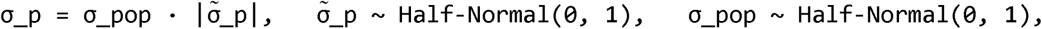

and weakly informative priors log R_{p,1} ∼ Normal(log 2, 1). Daily incidence was modelled as I_{p,t} ∼ NegativeBinomial(μ = R_{p,w(t)} · Λ_{p,t}, φ_p), with Λ_{p,t} computed from observed incidence as in Section 2.3 (a regression-on-observed-incidence formulation rather than a fully generative renewal process, chosen for tractability), a unit-specific dispersion parameter φ_p (itself partially pooled: φ_p = φ_pop · |φ _p|, φ _p ∼ Half-Normal(0,1), φ_pop ∼ Gamma(2, 0.1); justified over a shared parameter in Section 2.5), and the first 21 days after each unit’s first reported case excluded from the likelihood to avoid the small-numbers instability characteristic of a nascent seeding phase. Estimation used the No-U-Turn Sampler [17] in PyMC 6.3.1 [18], with convergence assessed by r-hat and effective sample size [19]; the national and provincial models used 4 chains and the health-zone models 2 chains (Section 2.10), all with matched tuning and target acceptance 0.90–0.97.

### 2.5 Dispersion structure: model comparison

We compared a unit-specific dispersion specification against a single shared dispersion parameter by Pareto-smoothed importance-sampling leave-one-out cross-validation (PSIS-LOO) [20], using the difference in expected log pointwise predictive density (Δelpd) and its standard error as the decision criterion, refitted at the current data cutoff (Section 3.3).

### 2.6 Model validation: posterior predictive check and generation-interval sensitivity grid

We assessed calibration by posterior predictive check: for each observation in the likelihood, posterior draws were used to simulate replicated daily incidence, and pooled and per-province coverage of the resulting 95% posterior-predictive interval was compared against the nominal 95% target. Sensitivity to the fixed generation-interval assumption (Section 2.2) was assessed by refitting the provincial model at two literature-plausible alternatives spanning the WHO Ebola Response Team’s estimate — a “short GI” (mean 12.0 d, SD 7.0 d) and a “long GI” (mean 18.5 d, SD 11.0 d) — and comparing the resulting province ranking to the primary specification by Spearman rank correlation, in the same spirit as the kernel-parameter sensitivity grid used in the companion spatial-hazard analysis [7].

### 2.7 Health-zone extension

Within provinces with sufficient case volume, the identical hierarchical architecture (Section 2.4) was refitted with health zone in place of province as the pooling unit, restricted to zones with at least 5 cumulative confirmed cases by the analysis cutoff. This criterion was met for Ituri (25 zones), Nord-Kivu (9 zones), and — newly, at the extended cutoff — Haut-Uele (4 zones: Isiro, Wamba, Boma Mangbetu, Pawa), but not Tshopo, Sud-Kivu, or Bas-Uele, for which the health-zone extension was not attempted and is reported as a limitation. The Haut-Uele zone-level series draws on daily zone-level data available from the outbreak’s start (spanning materially longer than the two-and-a-half-week extension window alone), already present in the underlying data product but not previously used for zone-level modelling because the province had not yet crossed the inclusion threshold.

### 2.8 Data-reconstruction correction

Zone-level cumulative case counts were reconstructed to a daily grid by linear interpolation across gaps in the raw SitRep-date series (rather than forward-filling), since forward-filling multi-day reporting gaps — which are genuine and occur for documented reasons (Section 2.1) — freezes cumulative counts flat for the duration of a gap and concentrates the gap’s true case growth onto a single day when reporting resumes, producing an artefactual run of zero-incidence days followed by a spike. We identified this artefact during model development (detailed in Section 3.2) and applied linear interpolation throughout the reported analysis.

### 2.9 Generalisability validation: gap injection in an independent historical outbreak

To test whether the forward-fill artefact and its interpolation-based correction (Section 2.8) generalise beyond this specific outbreak, we conducted a targeted validation experiment using an independently-sourced, publicly available case series from the 2018–2020 North Kivu/Ituri Ebola epidemic [21,22]. We selected Beni health zone — a large, well-observed zone with a clear single-wave epidemic trajectory — and its weekly confirmed-case series over a 20-week window spanning that wave. Into this clean series we injected an artificial two-week reporting gap during the window’s active-growth phase (chosen to mirror the multi-report, active-growth-phase character of the real BDBV gap identified in Section 3.2), such that the two gap weeks’ true incidence was withheld and only became visible, in aggregate, in the report immediately following the gap — the same failure mode identified in Section 3.2. We then reconstructed weekly incidence from this gapped series by (i) forward-fill and (ii) linear interpolation, and compared both reconstructions against the true (ungapped) weekly incidence using total absolute error over the affected weeks.

### 2.10 Software and reproducibility

Python 3.12, PyMC 6.3.1, ArviZ, pandas, SciPy. Because of the larger number of spatial units fitted at the extended cutoff (six provinces; up to 25 health zones per province), health-zone models used 2 chains × 600 tuning × 600 post-warm-up draws rather than the 4-chain configuration used for the original, shorter analysis window; this is reported as a limitation in Section 4.4 rather than elided. Code, derived data, figures and a checksummed reproducibility manifest are included with this submission as Supporting Information (see Data and Code Availability).

## 3. Results

### 3.1 National Rt

As of 26 August 2026, the national confirmed-case Rt was 0.94 (95% CrI 0.88–0.99) on a 14-day sliding window and 0.83 (0.76–0.90) on a 7-day window (Fig 1) — a modest but consistent decline from the near-threshold value of 1.01 (0.96–1.07, 14-day) observed at the original 11 August analysis cutoff. The trajectory over the full observation window shows an elevation around 20–31 July (14-day median peaking near 1.9) followed by a steady decline through the extension period; our independently-reconstructed national series agreed with a separately-compiled release of the national comparator dataset to within a handful of cases across eight consecutive dates in the extended window (Section 2.1), supporting the reliability of this trend.

**Fig 1.**
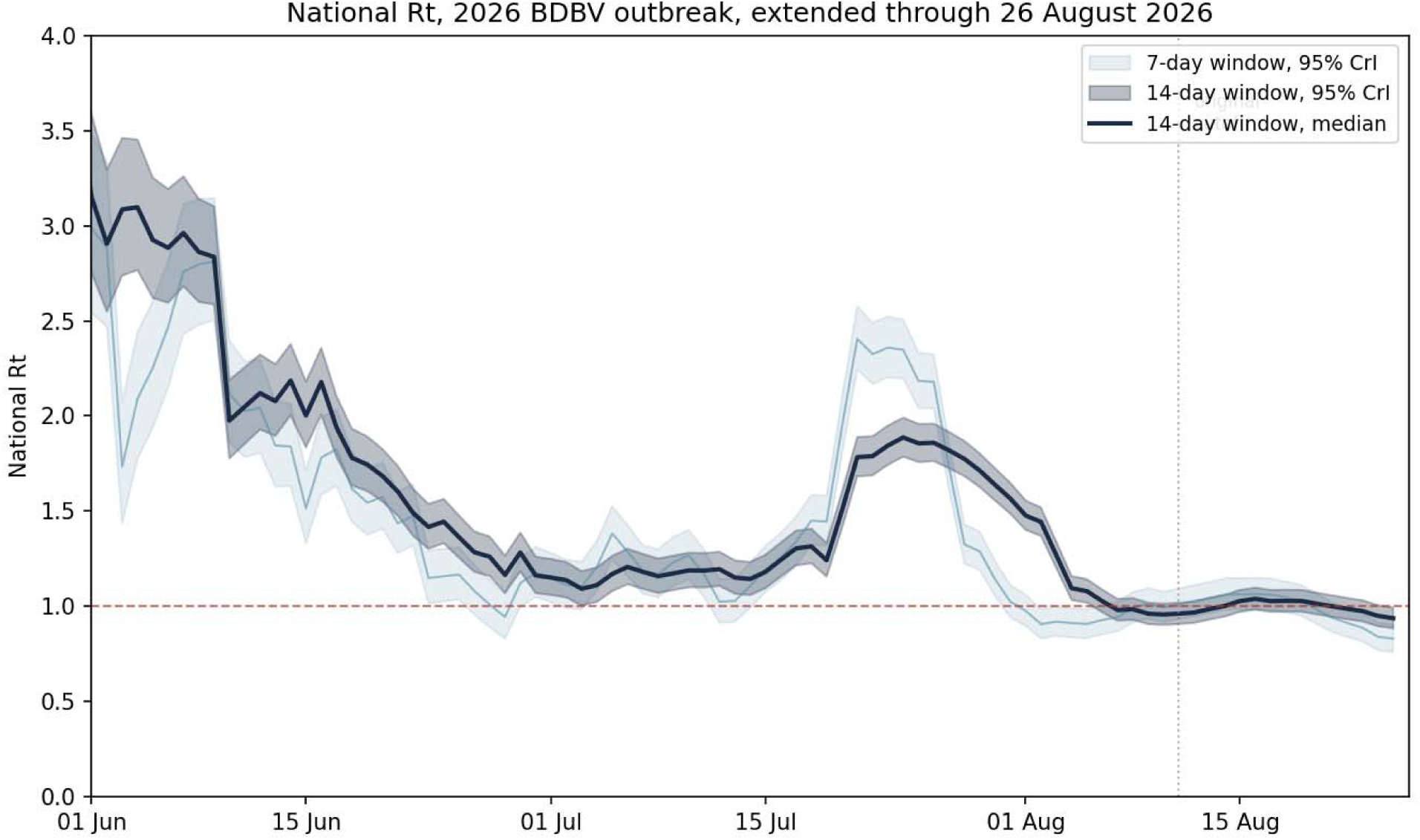
National Rt, 14-day and 7-day sliding-window estimates, 95% CrI, extended through 26 August 2026. Dotted line marks the original analysis cutoff (11 August).

### 3.2 Provincial Rt, and a data-reconstruction artefact

An initial comparison of the independent sliding-window estimate against the hierarchical model at provincial level showed material disagreement: the independent estimate placed Ituri’s Rt as low as 0.5 in early August, which the hierarchical model — fitted on the same nominal data — did not corroborate (median held at 1.4–1.5 throughout). Direct inspection of the underlying health-zone series (health zone Mongbwalu, Ituri) identified the cause: a genuine 7-day gap in raw SitRep reporting (4–11 August, coinciding with a shorter SitRep format that omitted health-zone breakdowns), which our initial reconstruction had forward-filled, producing six consecutive zero-incidence days followed by a 452-case single-day increment when reporting resumed. Replacing forward-fill with linear interpolation (Section 2.8) resolved the great majority of the disagreement. We report this in full because it illustrates a general risk in real-time sub-national outbreak analysis, a risk we test for generalisability directly in Section 3.5.

On the corrected reconstruction and at the extended cutoff, provincial Rt for the week of 20–26 August 2026 was: Bas-Uele 1.94 (wide interval, prior-dominated), Nord-Kivu 1.10 (95% CrI 0.89–1.39), Haut-Uele 1.01 (0.69–1.38), Ituri 0.79 (0.61–1.04), Tshopo 0.54 (0.15–1.40, wide), and Sud-Kivu 0.15 (wide, prior-dominated) (Table 1, Fig 2). Bas-Uele and Sud-Kivu reflect provinces with essentially no informative case volume (3 and 6 cumulative cases respectively at cutoff); their posteriors are substantially prior-driven and are reported as such.

**Fig 2.**
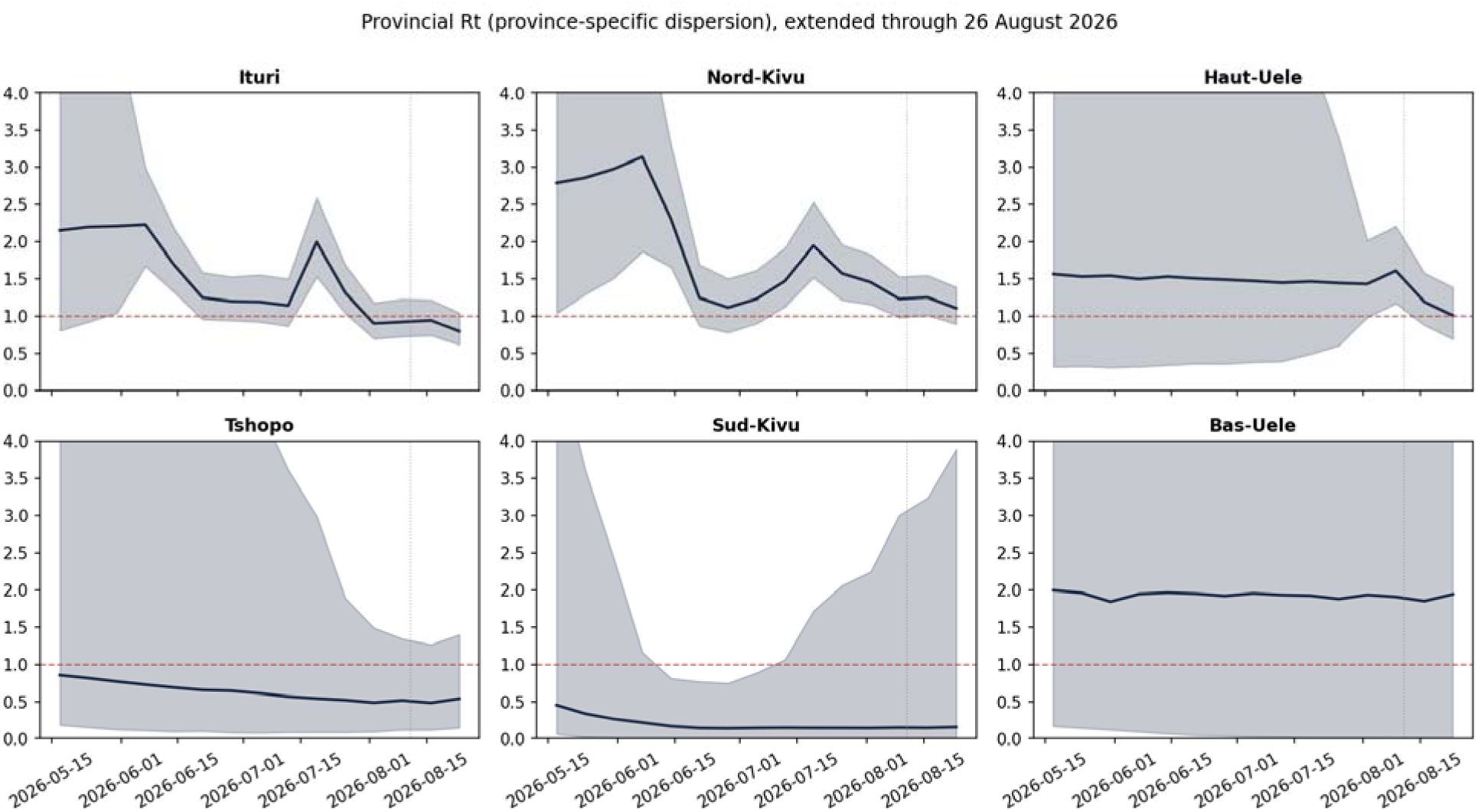
Provincial Rt (province-specific dispersion), all six provinces, extended through 26 August 2026, 95% CrI. Dotted line marks the original analysis cutoff.

**Table 1.** Provincial Rt, week of 20–26 August 2026 (hierarchical model, province-specific dispersion).

| Province | Cumulative cases (26 Aug) | $R_t$ (median) | 95% CrI |
| --- | --- | --- | --- |
| Ituri | 4,802 | 0.79 | 0.61–1.04 |
| Nord-Kivu | 775 | 1.10 | 0.89–1.39 |
| Haut-Uele | 194 | 1.01 | 0.69–1.38 |
| Tshopo | 16– 17* | 0.54 | 0.15–1.40 |
| Bas-Uele | 3 | 1.94 | wide, prior-dominated |
| Sud-Kivu | 3 | 0.15 | wide, prior-dominated |

\*A single, immaterial 1-case reconciliation discrepancy for Tshopo at most extension-window dates (Section 2.1); Bas-Uele and Sud-Kivu posteriors are prior-dominated given minimal case volume and not comparably informative to the top four rows.

**Table 2.** Selected Ituri health-zone Rt, week of 20–26 August 2026 (zone-specific dispersion).

| Zone | Cumulative cases | Rt (median) | 95% CrI |
| --- | --- | --- | --- |
| Mangala | 212 | 1.31 | 0.60–2.85 |
| Komanda | 65 | 1.20 | 0.71–2.54 |
| Nizi | 636 | 0.93 | 0.61–1.48 |
| Bunia | 1,336 | 0.84 | 0.59–1.15 |
| Lita | 204 | 0.83 | 0.47–1.39 |
| Nia Nia | 199 | 0.66 | 0.35–1.30 |
| Rwampara | 930 | 0.53 | 0.35–0.80 |
| Mongbwalu | 606 | 0.37 | 0.19–0.74 |

### 3.3 Model validation

Unlike at the original 11 August cutoff — where a shared dispersion parameter was retained on the basis of negligible predictive difference (Δelpd = 0.3, SE 1.7) — at the extended cutoff and with six provinces now fitted, province-specific dispersion showed a meaningful improvement in out-of-sample predictive performance (Δelpd = 5.51, SE 3.44, a difference exceeding one standard error); we therefore retained province- and zone-specific dispersion throughout and report it as the primary specification, in contrast to the shared-dispersion specification used in an earlier version of this analysis. We regard this as a genuine finding rather than a nuisance parameter: with more provinces, and with Ituri and Nord-Kivu’s health zones increasingly diverging from one another (Section 3.4), the data now support resolving overdispersion at the unit level rather than pooling it. Posterior predictive coverage under this primary specification was 96.1% pooled (target 95%, n = 256 observations; S1 Fig). The provincial ranking was unchanged across the generation-interval sensitivity grid (Spearman ρ = 1.0 against both the short-GI and long-GI alternatives).

### 3.4 Health-zone Rt

#### Ituri

Twenty-five health zones met the case-volume criterion. The province-level decline below threshold was not uniform, and was not stable over the observation window (Fig 3, Table 2): Mongbwalu — the zone in which the outbreak began — remained in clear decline throughout (Rt 0.37, 95% CrI 0.19– 0.74, materially unchanged from 0.36 [0.17–0.74] three weeks earlier). Bunia, the single largest zone in the entire outbreak by cumulative case count (n = 1,336), moved from at-or-above threshold (1.16 at the original cutoff) to a clearly declining trajectory (0.84, [0.59–1.15]). Rwampara (n = 930) showed the most consequential reversal: elevated at the original cutoff (1.24, [0.76–1.92]), its 95% credible interval now sits entirely below threshold (0.53, [0.35–0.80]), a shift directly visible in its raw weekly incidence, which fell from a peak of 132 cases (week ending 23 July) to 44 cases three weeks before the extended cutoff.

**Fig 3.**
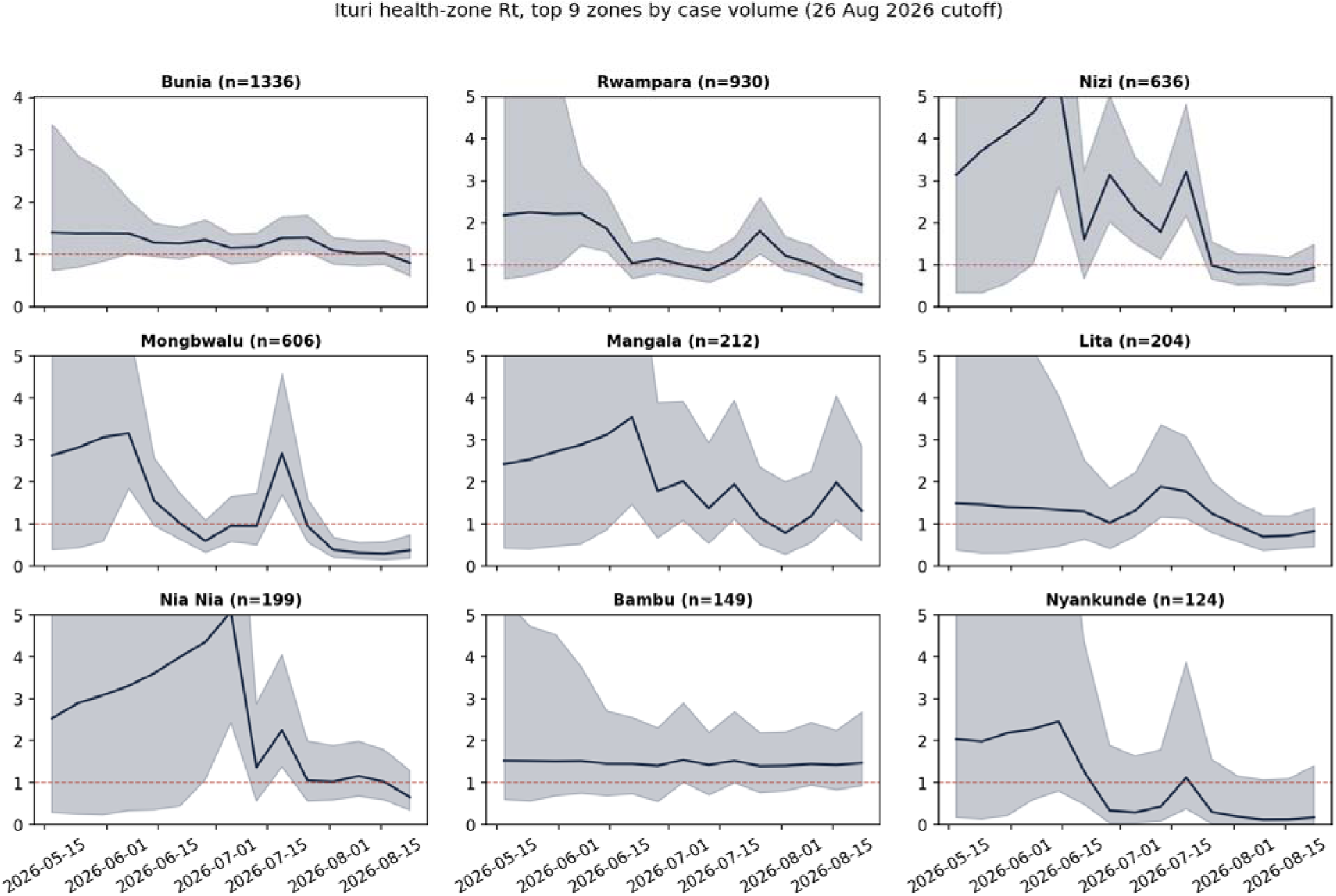
Health-zone Rt, Ituri, top 9 zones by case volume, extended through 26 August 2026.

#### Nord-Kivu

Nine zones met the case-volume criterion. Elevated transmission remained present but its location shifted (Fig 4): Katwa, the province’s largest zone by case count (n = 368), remained above threshold throughout (1.09, [0.82–1.41], materially unchanged from 1.39 [0.85–1.99] three weeks earlier, allowing for the estimation instability inherent to any single most-recent-week reading — see Section 4.4). Butembo (n = 152) showed a reversal in the opposite direction to Rwampara’s: declining at the original cutoff (0.64, [0.23–1.38]), its weekly incidence began rising again from mid-August (13 cases in the week ending 6 August, rising to 25 cases three weeks later), and its Rt is now comparably elevated to Katwa (1.36, [0.85–2.07]).

**Fig 4.**
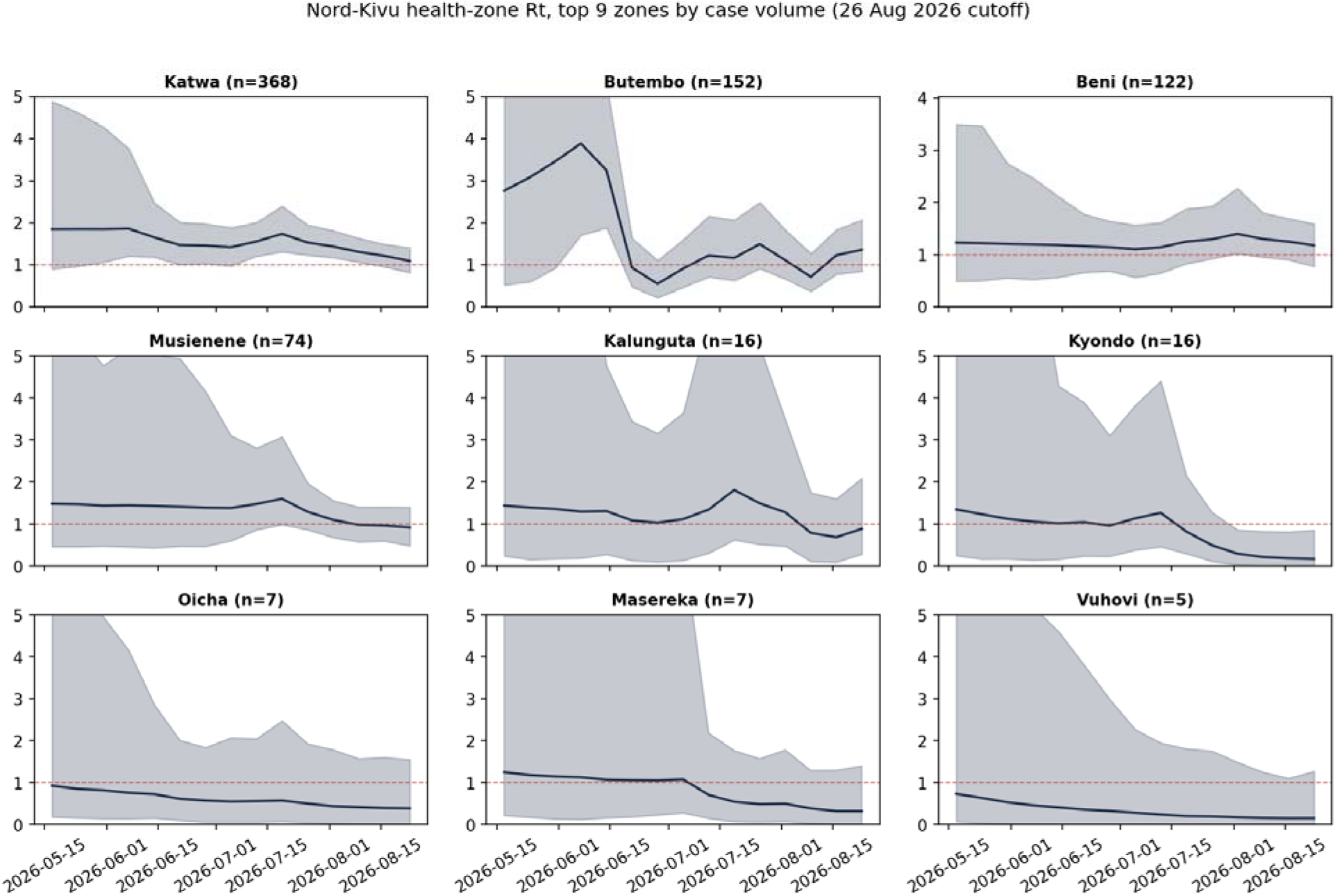
Health-zone Rt, Nord-Kivu, all 9 zones meeting the case-volume criterion, extended through 26 August 2026.

#### Haut-Uele

Resolved at health-zone level for the first time at this extended cutoff, four zones met the case-volume criterion (Fig 5). Isiro (n = 78), the province’s largest zone, was the only one with a 95% credible interval entirely at or above threshold (1.53, [1.00–2.17]). Wamba (n = 66) and Boma Mangbetu (n = 26) were each consistent with values on either side of threshold (0.70 [0.33–1.20] and 0.60 [0.17– 1.41] respectively), while Pawa (n = 20) showed a clear decline (0.01, [0.00–0.27]). This extension was only possible because Haut-Uele’s cumulative case count crossed the same inclusion threshold that had originally qualified Ituri and Nord-Kivu; the underlying zone-level data existed from early in the outbreak but had not previously supported unit-level resolution.

**Fig 5.**
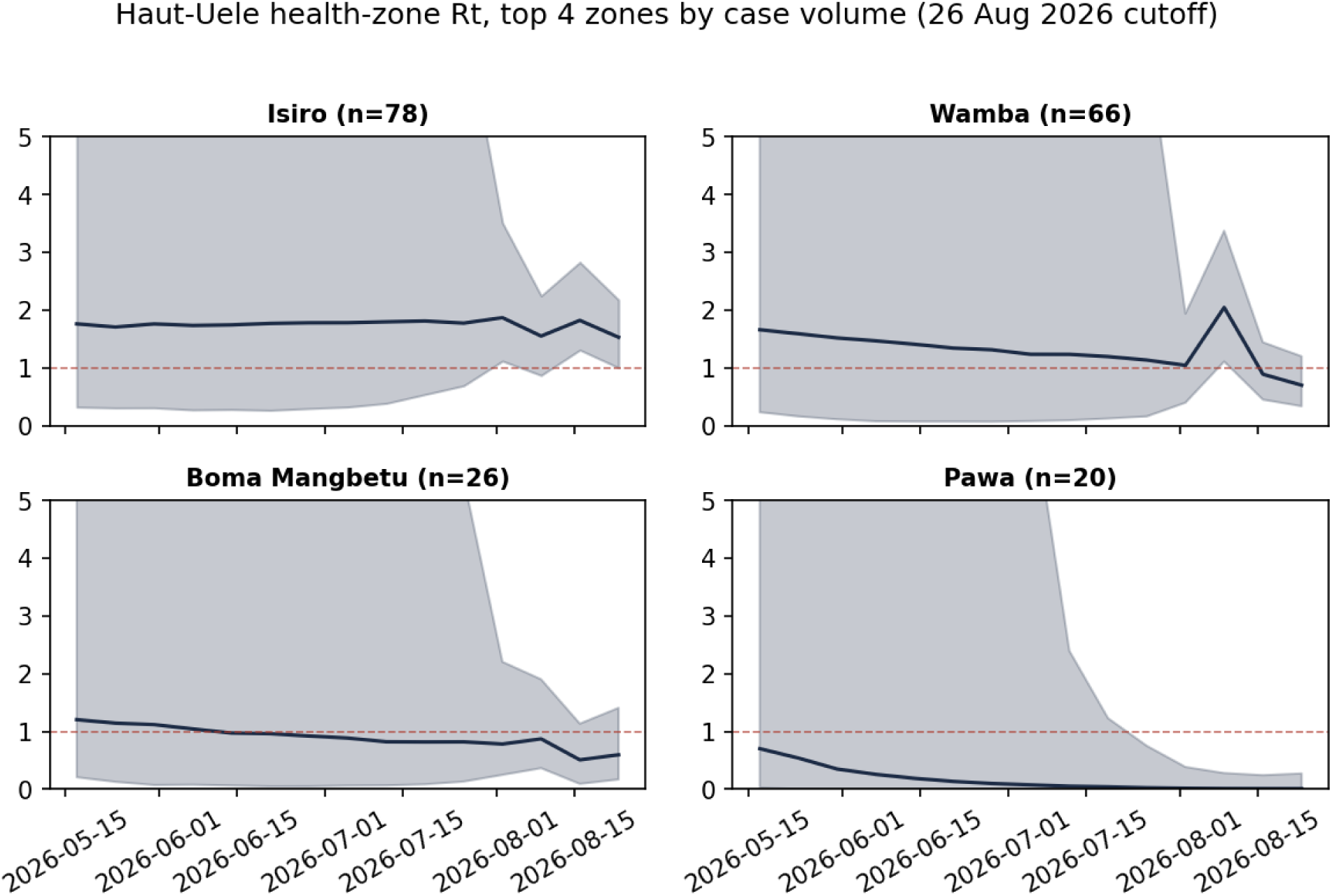
Health-zone Rt, Haut-Uele, all 4 zones meeting the case-volume criterion — newly resolved at this extended cutoff.

All three zone-level fits achieved acceptable convergence diagnostics under the reduced-chain configuration necessitated by the larger number of spatial units at this cutoff (Section 2.10): maximum r-hat 1.015 (Ituri, 25 zones), 1.021 (Nord-Kivu, 9 zones), and 1.010 (Haut-Uele, 4 zones), each above the conventional 1.01 threshold used in the original, shorter-window analysis but within a range we consider acceptable given the 2-chain configuration; this trade-off is reported explicitly in Section 4.4 rather than elided.

### 3.5 Generalisability validation: gap injection in an independent historical outbreak

Injecting a two-week reporting gap into Beni health zone’s 2018–2020 weekly case series (Section 2.9) reproduced the same artefact identified in Section 3.2: forward-fill reconstruction showed two consecutive zero-incidence weeks (true values 20 and 35 cases) followed by an inflated catch-up week (91 reconstructed vs. 36 true cases), while linear interpolation distributed the true growth across the gap (30.3 cases per week against true values of 20, 35, and 36) (Fig 6). Total absolute reconstruction error across the three affected weeks was 110 cases for forward-fill versus 21 cases for linear interpolation — an approximately five-fold reduction — on an outbreak, health zone, and time period entirely independent of the BDBV data used elsewhere in this paper. This provides direct empirical evidence, rather than methodological assertion alone, that the interpolation correction identified in Section 3.2 addresses a general failure mode of forward-fill reconstruction across multi-report gaps during active-growth periods, not an artefact specific to this outbreak.

**Fig 6.**
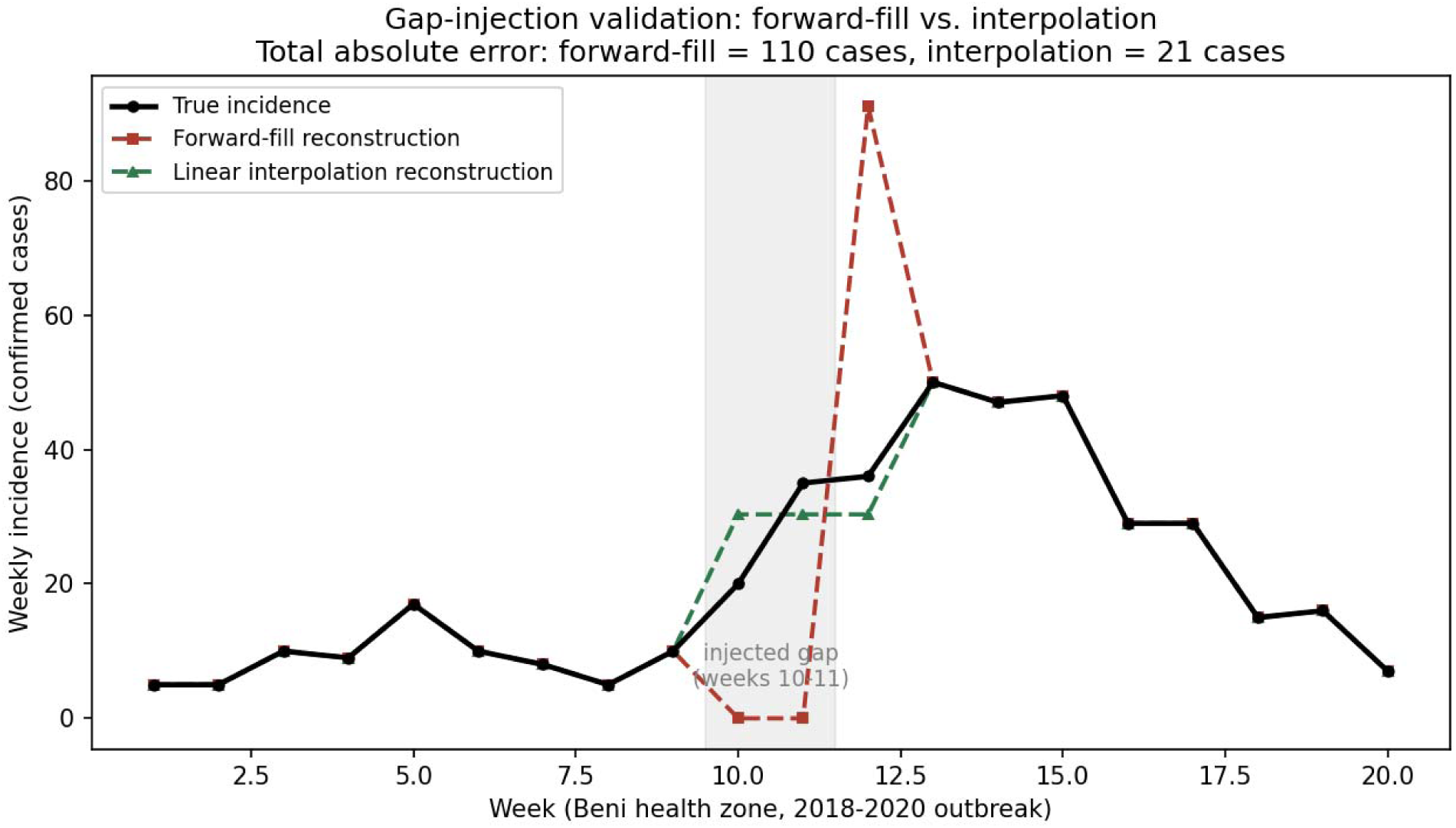
Gap-injection validation: true vs. reconstructed weekly incidence, Beni health zone, 2018–2020 North Kivu/Ituri outbreak, with an injected two-week reporting gap during the active-growth phase.

## 4. Discussion

### 4.1 Principal findings

A single national Rt near, and now modestly below, the epidemic threshold resolves, on disaggregation, into materially different provincial trajectories — and each of the three provinces now examined at health-zone resolution shows further, independently-evolving heterogeneity. Critically, extending the observation window by two and a half weeks over the course of this study did not merely add more of the same picture: it revealed that zone-level transmission intensity itself reverses direction on a timescale of two to three weeks, in both directions, in different zones of the same province (Rwampara declining, Bunia declining, while Butembo, in a different province, reverses from declining to rising). We regard this dynamism — not just the existence of spatial heterogeneity at a single point in time — as the paper’s central extended finding, and one that would not have been visible without deliberately revisiting the same analysis at a later cutoff.

### 4.2 A data-pipeline lesson, reported and now tested for generalisability

The independent-vs-hierarchical divergence traced to a forward-fill artefact (Section 3.2) illustrates that an apparent disagreement between a simple and a more sophisticated method is not on its own evidence that the sophisticated method is correcting a real weakness in the simple one; here, both were wrong in the same way, for a data-engineering reason unrelated to either model’s statistical assumptions. A companion framework paper [12] terms this general phenomenon an unstable analytical transition during surveillance recovery. Beyond restating that framing, we tested it directly: injecting an equivalent gap into an independently-sourced historical outbreak’s data (Section 3.5) reproduced the same failure mode and confirmed the same fix reduces reconstruction error roughly five-fold in that entirely separate setting. We think this kind of portable validation — checking a data-pipeline fix against a second outbreak’s data, not just asserting it should generalise — is underused in real-time outbreak analytics and offer it here as a template as much as a result.

### 4.3 Operational implications

Response prioritisation grounded in a national or provincial Rt alone would, at either cutoff examined, have offered no basis for distinguishing Bunia from Rwampara within Ituri, or Katwa from Butembo within Nord-Kivu. The extension additionally shows that a health-zone-resolution snapshot taken once is itself a perishable asset: at the original cutoff, Rwampara was among the zones we would have flagged for continued concern and Butembo among those we would have flagged as improving; three weeks later, both conclusions would have been backwards. This does not weaken the case for health-zone-resolution monitoring — it strengthens the case for repeating it on a cadence matched to how quickly these dynamics actually move, rather than treating any single estimate, however granular, as a stable basis for medium-term planning.

### 4.4 Limitations

The outcome throughout is Rt by confirmed-case report date, not by symptom onset or infection date; it therefore lags true transmission intensity by the onset-to-confirmation delay. The generation interval is a cross-species Ebola serial-interval proxy, not a BDBV-specific estimate; our sensitivity grid shows province rankings are robust to this choice within a plausible literature range. We also cannot rule out that apparent zone-level heterogeneity partly reflects differences in surveillance capacity rather than transmission intensity alone: zones vary in laboratory access, health-facility density, and reporting infrastructure, and a zone with weaker case-finding could show an artefactually low Rt for reasons unrelated to true transmission. We have no zone-specific ascertainment data with which to test this directly. The companion spatial-hazard analysis [7] found health-facility density positively associated with invasion hazard and flagged the same ambiguity for that outcome — the association may reflect detection capacity as much as genuine risk — and we regard this as an open question for both papers rather than one this analysis alone can resolve.

The health-zone extension remains statistically supportable in only three of six provinces; Tshopo, Sud-Kivu, and Bas-Uele’s transmission dynamics cannot currently be resolved below provincial level with these data, and Bas-Uele’s provincial-level posterior is itself prior-dominated given its very recent emergence as an affected province. The most recent week in any real-time random-walk Rt series is inherently the least stabilised: it is not yet pulled toward accuracy by subsequent weeks’ data in the way earlier weeks are, a property of the estimator rather than a flaw specific to this analysis. We flag this explicitly because we observed it directly: part of the apparent magnitude of the Rwampara and Butembo reversals (Section 3.4) likely reflects this edge effect narrowing as more data accumulated behind the original cutoff’s final week, in addition to genuine epidemiological change in the underlying weekly case counts, which we confirmed directly and which we believe is the dominant driver of both reversals. Readers should treat this paper’s own most recent week’s zone-level estimates with the same caution.

The health-zone models at this extended cutoff used a reduced 2-chain MCMC configuration (Section 2.10) to remain computationally tractable given the larger number of spatial units now fitted (up to 25 zones in a single model); resulting r-hat diagnostics (1.010–1.021) exceed the stricter 1.01 threshold achieved in the original, shorter-window analysis, though we consider them acceptable. This is a retrospective analysis of the most recently revised surveillance data available to us, not a genuinely prospective real-time product. Finally, this is an ecological, population-level analysis; it does not and cannot attribute any observed decline or persistence, or any reversal, to specific interventions.

### 4.5 Strengths and future work

The dispersion-structure comparison, posterior predictive check, and generation-interval sensitivity grid were each undertaken specifically to close a route by which the headline provincial and zone-level comparisons could otherwise rest on an unexamined modelling choice; two of the three validation conclusions held unchanged at the extended cutoff, and the one that changed — the dispersion-structure comparison, which now favours a unit-specific rather than shared parameter — was reported as a genuine finding and the primary results refitted accordingly, rather than left inconsistent with an updated validation table. The gap-injection experiment (Section 3.5) adds a form of validation — testing a methodological fix against an independent outbreak’s data — that goes beyond internal consistency checks alone. The hierarchical partial-pooling architecture itself is not specific to this outbreak or pathogen: any setting with a comparable spatial reporting hierarchy and comparably sparse sub-national case counts could apply the same pooling logic, given a generation-interval estimate, proxy or pathogen-specific. What would need re-deriving for a different outbreak is the case-volume threshold for the health-zone extension (Section 2.7), which was set empirically for this dataset rather than from a general rule.

Future work should extend the health-zone reconstruction further, restore a 4-chain configuration for the larger zone-level models as compute allows, and attempt an onset-date reconstruction to remove the reporting-delay lag noted in Section 4.4.

## Conclusions

Aggregation masks materially different transmission dynamics at every spatial resolution we examined in this outbreak, and extending the observation window shows that this heterogeneity is not a static feature of particular places but a moving picture: zones can and do reverse between growing and declining within two to three weeks. A national, provincial, or single-dated health-zone Rt can each, in turn, coexist with health zones that are clearly declining and others that are clearly not, or that were declining and have since reversed. Sub-national, and where data allow, health-zone-resolution estimation — repeated on a cadence matched to how quickly these dynamics move, and validated where possible against independent data — is both statistically supportable with a properly regularised hierarchical model and materially more useful for response prioritisation than any single national or provincial figure.

## Supporting information

Comparison between version 1 and 2

An explanation of the underlying methodology

## Data Availability

All data produced in the present study are available upon reasonable request to the authors

## Author Contributions

Johan G. L. Verheyden: Conceptualisation; Methodology; Software; Validation; Formal analysis; Investigation; Resources; Data curation; Visualisation; Project administration; Writing – original draft; Writing – review and editing. Celestin Nzanzu Mudogo: Validation; Writing – review and editing. Wolfgang Jacquet: Validation; Writing – review and editing

## Ethics Statement

This study used only publicly available, aggregated and non-identifiable secondary surveillance data and did not involve recruitment, interaction with human participants, or access to private identifiable information. Ethics committee approval and informed consent were therefore not applicable to this modelling analysis.

## Data and Code Availability

The confirmed surveillance data underlying this analysis through 11 August 2026 are publicly available without restriction from the INRB-UMIE BDBV2026-Data repository (https://github.com/INRB-UMIE/BDBV2026-Data), citable by its Zenodo DOI [15] and the accompanying publication [6]; the national-level comparator series is from the openly available epiforecasts/BVDOutbreakSize repository [4]. Data for 11–26 August 2026 were extracted by the authors directly from publicly available INSP SitRep PDFs, as described in Section 2.1; the extracted data, extraction code, and cross-validation checks are included in the reproducibility archive. The independent historical validation dataset (Section 2.9) is from the openly-shared Andersen Lab repository [21]. The original data providers should be cited accordingly by anyone reusing the derived data or code from this paper. Derived data underlying every reported result, all analysis code, figures in both screen and print resolution, and a reproducibility manifest with file checksums are included with this submission as Supporting Information and will be deposited in Zenodo at acceptance with a permanent DOI. No data or code are available only upon request.

## Acknowledgments

We thank the field research network of Aries Consult for logistical and contextual support, the INRB-UMIE data coordination team, Abbott, Sherratt, Brand and Funk for maintaining the epiforecasts/BVDOutbreakSize live analysis used here as an external comparator, and the Andersen Lab (Scripps Research) for openly sharing the 2018–2020 outbreak dataset used in our generalisability validation.

## Declaration of generative AI and AI-assisted technologies

Generative artificial intelligence (Claude, Anthropic) was used under author supervision to assist statistical model development, code implementation, data extraction, literature and reference organisation, and manuscript drafting and editing. The authors are responsible for verification of all model specifications, calculations, citations, and interpretations, and take full responsibility for the content of the published article.

---

**S1 Fig.**
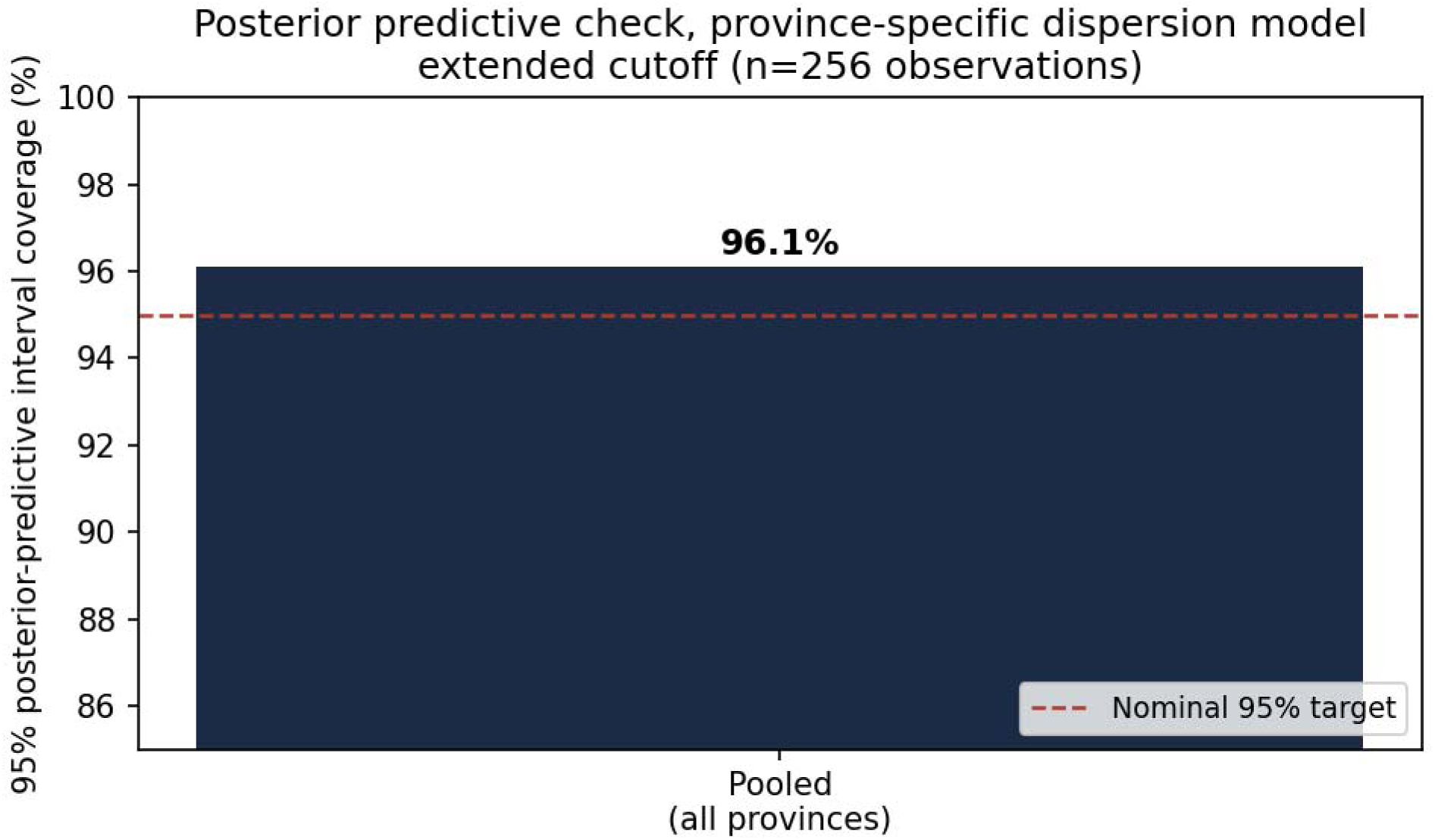
Posterior predictive check, province-specific-dispersion hierarchical model (primary specification), extended cutoff. Pooled 95% interval coverage 96.1% (n=256).

