## Supplementary material for "Sub-national heterogeneity in the time-varying reproduction number during the 2026 Bundibugyo virus disease outbreak in the Democratic Republic of the Congo: a hierarchical Bayesian analysis": Comparison between version 1 and 2

**Version Comparison and Decision Log**

Verheyden JGL & Nzanzu Mudogo C · Aries Consult · medRxiv 2026.08.19.26360792

### **1. Overview and Context**

This document records every substantive difference between the manuscript as submitted to PLOS Global Public Health (manuscript PGPH-D-26-03332, rejected after editorial review on 28 August 2026 without external peer review) and the revised version prepared for resubmission to PLOS Neglected Tropical Diseases. It is intended to serve three purposes: as an internal decision record, as a cover-letter annex explaining the revisions to the PLOS NTD editorial team, and as a reference for the reproducibility archive.

The rejection at PLOS GPH raised four specific concerns. Two were considered substantively valid and have been addressed by targeted text additions. One was considered a venue mismatch rather than a scientific problem. One remains contested but is addressed anyway to sharpen the methods section. Each is addressed in Section 2 below.

Separately from the GPH rejection, the analysis itself was revised in three consequential ways: the data cutoff was extended from 11 August to 26 August 2026; the dispersion structure of the model changed from shared to unit-specific following a repeated model comparison; and a new generalisability validation section was added. These are described in Sections 3–5.

| **Note** | The GPH submission was the result of a submission error — the manuscript was intended for PLOS NTD (for which the cover letter had already been written) but was accidentally transferred to PLOS GPH via the wrong dropdown during journal selection. This is noted here for completeness; the editorial feedback received is nonetheless addressed on its merits regardless of the original intent. |
| --- | --- |

### **2. Response to PLOS GPH Editorial Concerns**

The rejection letter (from Dr Katherine Demi Kokkinias, Staff Editor, PLOS Global Public Health, 28 August 2026) identified four concerns. Each is assessed below.

#### **2.1 Methods section: operationalisation and validation details**

**GPH concern:** "The Methods section lacks important information, such as operationalization feature of model and sufficient details regarding validation outcomes and the sensitivity analysis."

Assessment: Partly valid — as a venue mismatch artefact, partly genuine. The manuscript was written to a compressed word count calibrated for PLOS NTD conventions. The validation suite (PSIS-LOO dispersion comparison, posterior predictive check, GI sensitivity grid) was described in Sections 2.5–2.6 and results reported in Section 3.3 with specific numerical outcomes. However, GPH's editorial standard expects more expository prose explaining what each check tests and why, rather than the terse, equation-forward style appropriate for a specialist methods readership.

Action taken: The Methods sections (2.4–2.6) have been expanded with additional prose explanation of what the model's dispersion structure represents, what the PSIS-LOO comparison is testing, and what 'posterior predictive coverage' means operationally. The Results Section 3.3 now carries a more explicit narrative thread connecting each validation check to the modelling decision it tests. No numerical results were changed — only exposition was added.

**Additionally, the dispersion comparison finding itself changed substantively** between the GPH submission and this revision (see Section 4.3), so the validation section required a full rewrite regardless of the exposition concern.

#### **2.2 Health-zone definition and generalisability**

**GPH concern:** "Not enough description of how health zones were determined and if this method is generalizable to other outbreaks."

Assessment: Valid in two distinct respects. First, the term 'health zone' was used from its first appearance in the Introduction without ever being defined — a reasonable assumption for an outbreak-analytics or infectious-disease-epidemiology readership, but not for GPH's broader public health audience. Second, the question of whether the hierarchical partial-pooling approach generalises beyond this specific outbreak was never addressed in the manuscript; it was treated as self-evidently applicable, which is an assertion rather than an argument.

Action taken (definition): A parenthetical definition was added at the first use of 'health zone' in the Introduction: 'the sub-provincial administrative and surveillance unit around which DRC's health system is organised, broadly analogous to a health district.' This is a one-sentence addition that does not disrupt the paragraph's flow.

Action taken (generalisability): A new paragraph was added to Section 4.5 (Strengths and future work) stating plainly what would and would not transfer to a different outbreak: the hierarchical partial-pooling architecture generalises to any setting with a comparable spatial reporting hierarchy and sparse sub-national case counts; what would need re-deriving is the case-volume threshold for the health-zone extension (Section 2.7), which was set empirically for this dataset.

**Additionally, the gap-injection generalisability validation (Section 2.9 / 3.5), added for the revised version, directly addresses the generalisability question for the data-reconstruction methodology** by testing it on an independent historical outbreak's data — a more direct form of evidence than a methodological assertion.

#### **2.3 Reporting capacity and surveillance heterogeneity**

**GPH concern:** "Insufficient description of limitations around different reporting structures or capacity across health zones."

Assessment: Valid, and the sharpest of the four criticisms. The original Limitations section (Section 4.4) addressed report-date lag, fixed generation interval, shared dispersion, and retrospective design — but never asked whether zones with structurally weaker surveillance infrastructure might show artefactually low Rt independent of true transmission. A zone under-testing or under-reporting could appear to be declining when it is simply being less well observed. This is a genuine, unaddressed threat to the paper's core inference.

Action taken: A new paragraph was added to Section 4.4 naming the reporting-capacity confounder directly: 'We also cannot rule out that apparent zone-level heterogeneity partly reflects differences in surveillance capacity rather than transmission intensity alone: zones vary in laboratory access, health-facility density, and reporting infrastructure, and a zone with weaker case-finding could show an artefactually low Rt for reasons unrelated to true transmission.' The paragraph cross-references the companion spatial-hazard analysis [7], which independently flagged the same ambiguity for its own outcome (health-facility density may reflect detection capacity as much as genuine invasion risk), and frames both as an open question rather than a resolved one.

No analytical change was made — we have no zone-specific ascertainment data with which to test this directly. The limitation is reported honestly rather than addressed with an adjustment we cannot validate.

#### **2.4 Language and grammar**

**GPH concern:** "There are also concerns with language and grammar that we have identified and suggest thorough copyediting of the manuscript."

Assessment: Unable to verify or refute without specific examples, which were not provided in the rejection letter. No sentence was flagged, so this cannot be addressed point-by-point.

Action taken: A careful re-reading of the full manuscript was conducted; sentences were revised where passive constructions were unnecessarily convoluted, run-on sentences were split, and verb-tense consistency was enforced across the results section. The revised version is longer overall (due to the content additions in points 2.1–2.3 and the new validation section) but has been re-read in full by the corresponding author.

#### **2.5 Venue assessment**

PLOS Global Public Health's stated scope covers global health delivery, non-communicable diseases, race and health, mental health, humanitarian aid, and infectious diseases as one category among several. It is framed throughout around policy relevance, equity, and accessibility to 'health professionals, policy-makers, and local communities' — a broad practice/policy journal, not a specialist quantitative-methods venue.

Every specific editorial complaint in the rejection letter — wanting a plain-language definition of a health zone, wanting 'is this generalisable?' answered explicitly, wanting more expository walking-through of the validation — is exactly what happens when a technically dense Bayesian methods paper lands with a reviewer pool not built around hierarchical renewal modelling. This is not a criticism of PLOS GPH; it is a mismatch. The intended venue, PLOS NTD, serves a methodological infectious-disease readership for whom renewal-equation Rt estimation is standard background knowledge and for whom the validation suite described in Sections 2.5–2.6 needs no further unpacking.

The two valid criticisms (Sections 2.2 and 2.3) have been addressed regardless of venue, because a reviewer at PLOS NTD might make the same points and because both improve the paper on its merits.

### **3. Data Cutoff Extension (11 August → 26 August 2026)**

#### **3.1 Motivation and scope**

The GPH submission used a data cutoff of 11 August 2026 — the date through which the INRB-UMIE processed zone-level data product was current at the time of first analysis. By the time revisions were prepared, situation reports through 26 August 2026 (SitRep 104) were publicly available. Extending the cutoff was considered obligatory for a resubmission rather than optional: the outbreak was still active and evolving rapidly, and presenting two-week-old estimates as the current picture would have been misleading.

Extension required direct extraction from INSP SitRep PDFs for the period 11–26 August, since the processed data product had not yet incorporated this window. Nine SitReps (089, 093, 094, 096, 097, 099, 101, 102, 104) were parsed using a purpose-built table parser with cross-validation of every extracted province total against the same SitRep's own province-summary table. SitRep 094 (17 August) was excluded due to an anomalous table format that could not be parsed to province-level agreement.

#### **3.2 Key changes to results from the extended data**

| **Element** | **PLOS GPH submission (rejected)** | **Revised version (PLOS NTD resubmission)** | **Decision / rationale** |
| --- | --- | --- | --- |
| **Outbreak scope** | Five provinces (Ituri, Nord-Kivu, Haut-Uélé, Tshopo, Sud-Kivu).4,447 cumulative confirmed cases (11 Aug). | Six provinces (Bas-Uélé newly reported).5,794 cumulative confirmed cases (26 Aug). | Bas-Uélé added to provincial model (3 cases, prior-dominated posterior, reported as such). National count updated throughout. |
| **National Rt** | Rt = 1.01 (14-day, 95% CrI 0.96–1.07); 1.11 (7-day, 1.02–1.19). 'Near threshold, stable.' | Rt = 0.94 (14-day, 95% CrI 0.88–0.99); 0.83 (7-day, 0.76–0.90). Modest but consistent decline below threshold. | A real change in trajectory, not noise — visible in the 7-day estimate's shift from 1.11 to 0.83. Both abstract and results updated. |
| **Provincial Rt (Ituri)** | Rt = 0.91 (0.68–1.24), week of 6 Aug. | Rt = 0.79 (0.61–1.04), week of 20–26 Aug. | Continued decline, now with a tighter credible interval. Province-specific dispersion model used (see Section 4.3). |
| **Provincial Rt (Nord-Kivu)** | Rt = 1.23 (0.87–1.72), week of 6 Aug. | Rt = 1.10 (0.89–1.39), week of 20–26 Aug. | Above threshold but eased; interval tightened with more data. |
| **Provincial Rt (Haut-Uélé)** | Rt = 1.79 (1.24–2.67), week of 6 Aug. No zone-level model (case volume insufficient). | Rt = 1.01 (0.69–1.38), week of 20–26 Aug. Zone-level model now fitted (4 zones cross threshold). | Haut-Uélé crossed the zone-level case-volume threshold at the extended cutoff. Its zone-level data existed from early in the outbreak but had not been used. New Fig 5 and Table (Haut-Uélé zones) added. |
| **Ituri zone: Rwampara** | Rt = 1.24 (0.76–1.92), week of 6 Aug. Flagged as 'at or above threshold.' | Rt = 0.53 (0.35–0.80), week of 20–26 Aug. 95% CrI entirely below 1. | Genuine reversal confirmed in raw weekly incidence (132 cases wk ending 23 Jul → 44 cases three weeks before cutoff). The original paper's 'where to act' narrative required revision. |
| **Ituri zone: Bunia** | Rt = 1.16 (0.76–1.58), at or above threshold. | Rt = 0.84 (0.59–1.15), trending toward decline. | Real improvement. Bunia remains the largest zone in the entire outbreak (n=1,336) and keeps its featured position in the results. |
| **Nord-Kivu zone: Katwa** | Rt = 1.39 (0.85–1.99). Still growing. | Rt = 1.09 (0.82–1.41). Above threshold but eased. | Persistent elevation confirmed; remains a headline finding. |
| **Nord-Kivu zone: Butembo** | Rt = 0.64 (0.23–1.38). 'Already declining.' | Rt = 1.36 (0.85–2.07). Reversed to elevated trajectory. | Genuine reversal confirmed in raw incidence (13 cases wk ending 6 Aug → 25 cases three weeks later). Previous framing as 'success story' counterposed with Katwa required full revision in Results and Discussion. |
| **Figures** | 4 figures + S1 Fig. | 6 figures + S1 Fig. New: Fig 5 (Haut-Uélé zone Rt), Fig 6 (gap-injection validation). Updated: Fig 1–4 with extended data. | Added to reflect new province zone extension (Fig 5) and new validation section (Fig 6). |

#### **3.3 The 'most recent week' estimation instability**

Part of the apparent magnitude of both reversals (Rwampara and Butembo) reflects a structural property of real-time random-walk Rt estimation, not purely epidemiological change: the most recent week's estimate is always the least anchored, because it has no subsequent weeks pulling it toward stability. As more data accumulate behind the original cutoff's final week, that week's estimate naturally stabilises. This means some of what appeared as a definitive 'at threshold' reading for Rwampara on 11 August was partly estimation uncertainty that only resolved once 14–19 more days of incidence data were available. The real-time edge effect is now flagged explicitly in Section 4.4, and readers are advised to treat this paper's own most recent week's zone-level estimates with the same caution.

### **4. Modelling Changes**

#### **4.1 Dispersion structure: shared → unit-specific**

This is the most consequential analytical change between the two versions and the one where the decision is most worth documenting in detail.

##### **Original specification (GPH submission)**

A single shared dispersion parameter φ was used across all provinces (and zones within a province). This was chosen at the 11 August cutoff because PSIS-LOO showed negligible predictive difference between shared and province-specific dispersion: Δelpd = 0.3, SE 1.7 — well within one standard error. The shared specification was simpler, faster, and the data did not support preferring the more complex alternative.

##### **Revised specification**

Province- and zone-specific dispersion is used throughout. At the 26 August cutoff, with six provinces fitted, PSIS-LOO now shows: Δelpd = 5.51, SE 3.44 — a difference exceeding one standard error, meaning province-specific dispersion achieves meaningfully better out-of-sample predictive accuracy. The data now support the more complex specification.

##### **Why the conclusion changed**

Two factors drove the change. First, the number of provinces increased from five to six (Bas-Uélé added), giving the LOO comparison more statistical power to detect genuine differences in overdispersion. Second, and more substantively, the zones themselves had diverged further by late August: Rwampara's decline and Butembo's reversal within the same outbreak mean the assumption that all zones share a common overdispersion is increasingly implausible. The comparison reflects a genuine change in what the data support, not a change in preference.

| **Decision** | Province- and zone-specific dispersion is retained as the primary specification and reported as a genuine finding, not a nuisance parameter. The original shared-dispersion comparison is reported transparently (Δelpd = 0.3 at the earlier cutoff) to show the change in the data's evidentiary position, not hidden. Both the LOO result and the methodological reasoning are in Section 3.3 of the revised manuscript. |
| --- | --- |

##### **Effect on headline numbers**

Provincial point estimates (posterior medians) shifted negligibly — the largest change was 0.05 Rt units (Bas-Uélé, a prior-dominated province). Dispersion structure primarily affects uncertainty width and calibration, not where the posterior median sits. The zone-level point estimates similarly shifted by less than 0.05 units across all key zones. The Rwampara and Butembo reversals are robust to the dispersion specification — they appear under both shared and zone-specific dispersion, confirming they are driven by the underlying incidence data, not by the model choice.

#### **4.2 Chain configuration for zone-level models**

The original zone-level models (Ituri 24 zones, Nord-Kivu 8 zones) used 4 chains × 800 post-warmup draws. The revised models (Ituri 25 zones, Nord-Kivu 9 zones, Haut-Uélé 4 zones) use 2 chains × 600 draws due to the increased number of spatial units and the compute constraints of this analysis environment.

Maximum r-hat: 1.015 (Ituri), 1.021 (Nord-Kivu), 1.010 (Haut-Uélé). These exceed the conventional 1.01 threshold achieved in the original analysis. This is reported explicitly in Section 4.4 of the revised manuscript as a limitation, not elided. Qualitative conclusions are unchanged and the posteriors are considered reliable for the purposes of this analysis, but replication with a 4-chain configuration is flagged as a priority for future work.

#### **4.3 Posterior predictive coverage**

| **Element** | **PLOS GPH submission (rejected)** | **Revised version (PLOS NTD resubmission)** | **Decision / rationale** |
| --- | --- | --- | --- |
| **Primary model** | Shared dispersion. 95.0% pooled PPC coverage (n=200). Per-province range: 93–97%. | Province-specific dispersion (primary). 96.1% pooled PPC coverage (n=256). Note: per-province breakdown not separately saved; pooled figure only reported. | Coverage slightly improved with the new specification and extended data. The figure in S1 now shows pooled coverage only, without a fabricated per-province breakdown. |
| **GI sensitivity grid** | Spearman ρ=1.0 for province ranking across main/short/long GI variants (5 provinces). | Spearman ρ=1.0 for province ranking across main/short/long GI variants (6 provinces, including Bas-Uélé). | Conclusion unchanged. Grid rerun at extended cutoff with province-specific dispersion to maintain consistency with primary specification. |
| **LOO comparison** | Δelpd = 0.3 ± 1.7. Shared dispersion retained (negligible difference). | Δelpd = 5.51 ± 3.44. Province-specific dispersion adopted as primary (exceeds 1 SE). Both results reported. | A genuine methodological finding, not a nuisance: the data now support the more expressive model. Reported transparently in Section 3.3. |

### **5. New Section: Generalisability Validation (Section 2.9 / 3.5)**

#### **5.1 Motivation**

The PLOS GPH rejection explicitly asked about generalisability. The data-reconstruction correction (linear interpolation over forward-fill for multi-day SitRep gaps) was described in the GPH submission as a general principle, but its claimed generalisability rested on methodological assertion rather than evidence. A reviewer at any venue might ask: 'Does this specific fix actually work on a different outbreak's data, or did you only demonstrate it for the specific case where you found the problem?'

The gap-injection validation was designed to answer this directly: take the correction, apply it to a completely independent historical outbreak's case series (Beni health zone, 2018–2020 North Kivu/Ituri Ebola epidemic, Andersen Lab / Scripps Research), inject an artificial gap matching the structural pattern of the real BDBV gap, and quantify whether forward-fill or interpolation is more accurate.

#### **5.2 Design**

Beni health zone was selected because it is large, well-observed, and has a clear single-wave epidemic trajectory with no ambiguity about the true trajectory. A 20-week window spanning the main growth phase was used. A two-week reporting gap was injected during the active-growth phase (weeks 9–10 of the window, true incidence 20 and 35 cases), mirroring the BDBV gap: multi-report, active-growth phase, data appearing in aggregate in the first post-gap report. Both forward-fill and linear interpolation were used to reconstruct weekly incidence from the gapped series.

#### **5.3 Result**

Total absolute reconstruction error: 110 cases for forward-fill, 21 cases for linear interpolation — a 5.3-fold reduction. The forward-fill produced two consecutive zero-incidence weeks followed by an inflated catch-up week (91 reconstructed vs. 36 true cases), reproducing exactly the same failure mode identified in Section 3.2 for the BDBV data.

| **Key finding** | The correction generalises. The same failure mode (forward-fill during an active-growth reporting gap) and the same fix (linear interpolation) produce a five-fold error reduction on an outbreak, health zone, and time period entirely independent of the BDBV data. This converts a methodological assertion into an empirical claim with evidence. |
| --- | --- |

#### **5.4 Data source and attribution**

The historical dataset is openly available from the Andersen Lab (Scripps Research) GitHub repository (https://github.com/andersen-lab/ebola-drc-epidemiology) and is cited in the revised manuscript as references [21] and [22]. The dataset is also included in the reproducibility archive (data/raw/andersen_lab_2018_2020_ebola_drc.csv). The gap-injection analysis script is script 07 in the reproducibility archive.

### **6. Structural and Framing Changes**

#### **6.1 Framing shift: heterogeneity as dynamic, not static**

The GPH submission framed the paper's central contribution as 'aggregation hides spatial heterogeneity' — a cross-sectional argument about the value of sub-national resolution at a single point in time. The revised version adds a second, temporally-oriented argument: the heterogeneity is not static, and zone-level transmission intensity itself reverses direction on a timescale of two to three weeks. This argument was available only once the cutoff was extended and the Rwampara/Butembo reversals became visible. It is now explicitly named in Section 4.1 as the paper's 'central extended finding' and appears in the Abstract, Author Summary, and Conclusions.

#### **6.2 Abstract: substantially rewritten**

The abstract required full revision to reflect: the changed national Rt finding; the extended cutoff; the six-province scope; the Haut-Uélé zone extension; the Rwampara and Butembo reversals; the changed dispersion specification; the gap-injection validation; and the revised PPC coverage figure. Essentially every numerical result in the abstract changed. Length is similar to the GPH version.

#### **6.3 Author Summary: updated**

Updated to reflect the Rwampara reversal (a zone that 'was still growing three weeks ago has since turned around') and Butembo ('one town that had been declining has since reversed'), the gap-injection validation, and the corrected national trajectory.

#### **6.4 Word count**

| **Element** | **PLOS GPH submission (rejected)** | **Revised version (PLOS NTD resubmission)** | **Decision / rationale** |
| --- | --- | --- | --- |
| **Word count** | ~5,600 words | ~7,109 words | Increase reflects: new methods sections (2.7 Haut-Uélé extension, 2.9 gap injection); new results sections (3.4 Haut-Uélé, 3.5 gap injection); expanded limitations; expanded strengths. PLOS NTD has no hard word limit. |
| **Tables** | 2 (provincial Rt, Ituri zone Rt) | 3 (provincial Rt, Ituri zone Rt, new Haut-Uélé zone Rt) | Haut-Uélé zone table added as a parallel to Tables 1 and 2. |
| **Figures** | 4 manuscript + S1 | 6 manuscript + S1 | Fig 5 (Haut-Uélé zone trajectories) and Fig 6 (gap-injection validation) added. |
| **References** | 20 | 22 | References [21] (Andersen Lab dataset) and [22] (Aruna et al. MMWR 2019) added for gap-injection validation attribution. |

### **7. What Did Not Change**

The following are explicitly documented as unchanged, to make clear they were re-examined and not inadvertently altered:

- Model architecture: the hierarchical random-walk renewal equation, partial pooling structure, NUTS sampler, PyMC 6.3.1 implementation, and the first-21-days exclusion criterion are all identical between versions.
- Generation interval: unchanged (WHO Ebola Response Team 2014 proxy, mean 15.3d, SD 9.3d). Sensitivity grid rerun at extended cutoff; conclusion (ρ=1.0) unchanged.
- Health-zone inclusion criterion: ≥5 cumulative confirmed cases, unchanged. The new provinces that cross this threshold do so because of their own accumulated data, not because the threshold was relaxed.
- Companion-paper non-overlap statements: all seven companion papers were re-read at the extended cutoff to confirm the non-overlap assessments in Section 1.1 remain accurate. No new overlap was identified.
- Data attribution and open-data statements: unchanged and verified against current repository states.
- Author contributions, ethics statement, competing interests: unchanged.
- Cover letter: the PLOS NTD cover letter was written before the GPH submission and required only updating the medRxiv DOI (now known: 10.64898/2026.08.19.26360792) and a brief addition noting the editorial history. It did not require revision of the scope argument, which was written for NTD from the start.

### **8. Outstanding Items for Submission**

| **Element** | **PLOS GPH submission (rejected)** | **Revised version (PLOS NTD resubmission)** | **Decision / rationale** |
| --- | --- | --- | --- |
| **medRxiv v2** | N/A | A v2 preprint update is warranted given the substantive changes to results and the new validation section. The local docx is updated; posting is a separate decision. | Recommend posting v2 before or simultaneously with PLOS NTD submission to give the WHO Collaboratory audience and other readers the current numbers. |
| **WHO Collaboratory deck** | N/A | The 8-slide English presentation was built using the 11 August data and now mismatches the paper. Key numbers affected: national Rt, Rwampara/Butembo framing, Haut-Uélé. | Should be updated before any further use. Low priority if the 27 August presentation has already concluded. |
| **Operational poster** | N/A | The French-language operational poster (A1 SVG) was built with the 11 August choropleth and still references 'DOI à ajouter.' | DOI and data cutoff should be updated. Lower priority than the manuscript itself. |
| **PLOS NTD cover letter date** | N/A | The cover letter requires a submission date before filing. | Add date before submitting. |
| **Zenodo deposit** | N/A | Reproducibility archive is ready (BDBV_Rt_reproducibility_archive.zip); Zenodo deposit should happen at acceptance, per Data Availability statement. | No action needed at submission; flagged for post-acceptance. |

*Document prepared by Johan G. L. Verheyden · Aries Consult · 2026-08-30*
