## Supplementary material for "Sub-national heterogeneity in the time-varying reproduction number during the 2026 Bundibugyo virus disease outbreak in the Democratic Republic of the Congo: a hierarchical Bayesian analysis": An explanation of the underlying methodology

**Rt, Explained the Feynman Way**

Verheyden JGL, Mudogo CN & Jacquet W. · Aries Consult · medRxiv 2026.08.19.26360792

**A promise before we start**

Every single piece of math in the paper is something you can actually understand — not just “accept because a computer said so.” None of it is magic. Some of it is clever, but clever things can still be explained simply. That's the whole idea of this document: we're going to go slowly, use real numbers from the actual outbreak, and build every idea out of things you already know — counting, fractions, guessing games, and a little bit of “if this, then that.”

**How to use this document**

Each part below matches a section of the real paper (I'll tell you which one). Read them in order the first time — each part uses ideas from the one before it, the same way you can't understand fractions before you understand division. After that, you can jump around and use it as a reference.

One more thing, and it matters: simplifying HOW something is explained is not the same as making the thing itself less true. Every number in this document is real, taken directly from the paper. I'm not going to round off the hard parts — I'm going to build up to them.

**Part 0 · The One Idea Everything Else Is Built On**

*Matches: Section 1, Introduction*

Imagine you tell a secret to one friend. If that friend tells it to two more friends, and each of THOSE friends tells it to two more, the secret spreads fast — it doubles every time it gets passed on.

Now imagine instead that, on average, your friends only tell HALF a person the secret — meaning sometimes they tell someone, sometimes they don't bother. The secret dies out. Fewer and fewer people know it each round, until nobody's telling it anymore.

That's it. That's Rt. “R” stands for reproduction number. It just means: **on average, how many new people does one infected person go on to infect?**

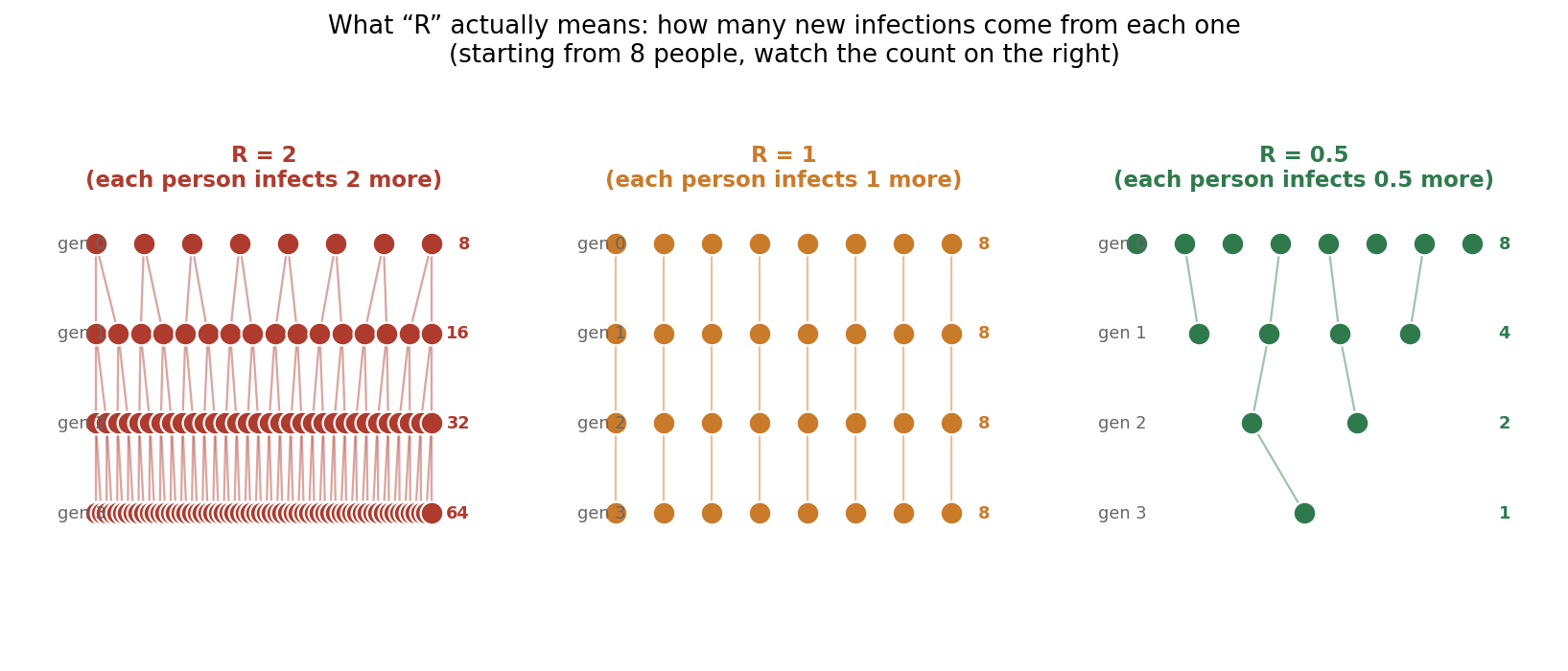

*Figure A — Three outbreaks that start identically and end completely differently, because of one number.*

Look at that picture. All three groups start with the same 8 people. That's the ONLY thing that's the same. Everything else — whether the outbreak explodes to 64 people or shrinks to a single person — comes down entirely to whether R is bigger than 1, exactly 1, or smaller than 1.

**Why “1” is the magic number**

If R > 1, each generation is bigger than the last — growing, like the red tree. If R = 1, each generation replaces itself exactly — flat, like the middle tree. If R < 1, each generation is smaller — shrinking toward zero, like the green tree. This is the single most important number in the whole paper. Everything else exists to answer one question, over and over, at different places and times: is R above 1 or below 1, right here, right now?

The paper's headline number, as of 26 August 2026, is national Rt = 0.94. That means: right now, on average, every 100 infected people in the whole country are currently infecting about 94 more. Slightly shrinking — but only just.

Here's the problem the whole rest of the paper is about: that “94 out of 100” is an AVERAGE across the entire country. Averages can hide really different realities underneath them. If one town's outbreak is roaring (say, R = 1.5) and another town's is dying out (R = 0.5), the average could easily land right around 1 — looking calm on the surface while hiding real trouble underneath. Finding out where that trouble actually is, is what the rest of this paper does.

**The one goal behind every choice you're about to read**

Before Part 1, here's something worth holding onto for the rest of this document. Every single mathematical and statistical choice described from here on — every formula, every distribution, every validation check — exists to serve exactly one goal:

**The goal, in one sentence**

Detect real differences between places, and real changes over time, without being fooled by noise — and be honest about how confident to be in every single number reported.

That's it. That's the whole job. Watch for this as you read: almost every time the paper does something that looks like extra complexity for its own sake, it's actually protecting one of those three things — catching real differences, not being fooled by noise, or being honest about confidence. From here on, each Part includes a dashed purple box explaining exactly which part of that job a given choice is doing, and what would go wrong without it.

**Part 1 · Turning a Scoreboard Into “Cases Per Day”**

*Matches: Section 2.1 and 2.8, Data sources and data-reconstruction correction*

Health reports don't usually say “14 new cases today.” They say “4,802 cases so far.” That's a running total — like a basketball scoreboard that only ever goes up. To find out how many NEW cases happened on one particular day, you do the simplest possible math: subtract yesterday's total from today's total.

new cases today = total today − total yesterday

That works perfectly... as long as you get a report every single day. But real outbreaks don't work that way. Sometimes a health worker can't file a report for a week because of bad roads, or a form got redesigned, or a printer broke. What happens then?

**The trap: freezing the score**

Imagine your team's score is 10 points on Monday. Nobody updates the scoreboard for a week. Then on the next Monday, it suddenly reads 45. The laziest way to fill in the missing week is to assume nothing happened until the last moment — score stayed at 10 all week, then BOOM, all 35 new points landed on that one final day.

That's called forward-fill, and it's exactly what happened by accident early in this project. A real 7-day gap in reporting (4–11 August, in a health zone called Mongbwalu) got treated this way — which invented six days that LOOKED like nothing was happening, followed by a single day that looked like an explosion of 452 new cases. Neither of those things was true. The real cases were spread out across the whole week; the data just didn't say so yet.

**The fix: connect the dots**

If you know the score was 10 on Monday and 45 the following Monday, and you have no other information, your best honest guess for the days in between is a straight line from 10 up to 45 — a little bit more each day. That's called linear interpolation. It's the same idea as: if you were 140 cm tall on your 10th birthday and 152 cm tall on your 12th birthday, your best guess for your height on your 11th birthday is right in the middle, about 146 cm — not “still 140 until the last second, then a sudden jump.”

**WHY THIS CHOICE, NOT SOMETHING SIMPLER?**

You might ask: does it really matter which way you fill a gap, as long as the running total ends up right by the next report? It matters enormously, because Rt estimation (Part 2) doesn't look at running totals — it looks at the SHAPE of day-to-day case counts, comparing recent days to a weighted history of past days. A fake zero-then-spike pattern doesn't just blur the picture a little; it directly poisons the exact quantity being measured, at the exact moment accuracy matters most. This choice protects “not being fooled by noise” — except here the noise isn't in the data, it's accidentally created by how the data was PROCESSED. That's a subtler danger than noisy data itself, because it can look perfectly clean and confident right up until you check it against the truth (which is exactly what Part 6 does).

This sounds almost too simple to be worth a whole section of a scientific paper — but it turned out to matter enormously. Two DIFFERENT ways of estimating Rt (a simple one and a fancier one, both described later) disagreed with each other early in this project, and it looked like the fancier method was somehow “correcting” the simple one. It wasn't. Both methods were being fed the same bad, frozen-then-spiking data. Once the interpolation fix was applied, the disagreement mostly vanished. The lesson: sometimes when two smart methods disagree, the problem isn't either method — it's the data they were both trusting.

Part 6 of this document describes a real experiment the authors ran to check that this fix genuinely works, not just on this outbreak, but on a completely different one too.

**Part 2 · The Simple Way to Guess Rt**

*Matches: Section 2.2 and 2.3, Generation interval and the sliding-window estimator*

**Idea 1: infections have an echo, not an instant effect**

When someone catches BDBV, they don't immediately go on to infect somebody else that same day. There's a delay: they need to develop symptoms, come into contact with other people, and pass it on. For this specific virus, the paper uses an estimate that this typical delay averages about 15.3 days, though it varies a lot from person to person (some pass it on sooner, some later).

This delay is called the generation interval. Think of it like an echo in a canyon: you shout (get infected), and the echo (you infecting someone else) doesn't come back instantly — it takes time, and it's loudest around a typical delay, fading out on either side.

**Idea 2: today's new cases are mostly explained by cases from about two weeks ago**

If today's new infections were mostly CAUSED by people who got infected roughly 15 days ago (because that's the typical delay before passing it on), then you can build a prediction: add up all the recent daily case counts, but give MORE weight to days about 15 days back, and LESS weight to days that are much closer or much further away. The paper calls this weighted sum the “infectiousness” on a given day, written as the Greek letter Lambda (Λ).

**WHY THIS CHOICE, NOT SOMETHING SIMPLER?**

Why not just watch whether daily case counts are going up or down, and skip the “echo” idea entirely? Because rising case counts don't necessarily mean transmission is speeding up RIGHT NOW — they could simply be the delayed consequence of a lot of transmission that already happened two weeks ago, still working its way through the reporting pipeline. Without the generation-interval echo, you'd be measuring the PAST, but mistaking it for the PRESENT. The whole point of Rt is to isolate current transmission risk, adjusted for how many currently-infectious “seeds” are already in the population — that adjustment is exactly what Λ provides, and it's the difference between a genuinely useful early-warning number and a number that's always a bit late to the story.

Λ (today) = a weighted echo of new cases from the past(most weight around 15 days ago, less weight further out)

**Idea 3: compare what actually happened to what the echo predicted**

Here's the actual heart of it. If the number of new cases we're SEEING today is about the same as what the echo (Λ) predicts, transmission is holding steady — Rt ≈ 1. If we're seeing MORE new cases than the echo predicts, something is amplifying transmission — Rt > 1. If we're seeing FEWER, transmission is fizzling out — Rt < 1.

Rt ≈ (new cases actually happening) / (Λ, what the echo predicted)

**Idea 4: don't give one number, give a whole range of honest guesses**

Here's a subtlety that matters a lot for small places. Imagine you flip a coin 5 times and get 4 heads. Does that mean the coin is biased to land heads 80% of the time? Obviously not — 5 flips is nowhere near enough to be sure. But if you flipped it 5,000 times and got 4,000 heads, NOW you'd be pretty confident it really is biased.

The same logic applies to counting infections. A province with only 3 total cases (like Sud-Kivu) can look wildly “growing” or “shrinking” from day to day just by random chance — the same way 5 coin flips can look biased just by luck. A province with 4,802 cases (like Ituri) has enough data that random noise mostly cancels out.

**What a “Bayesian” approach actually does**

Instead of reporting one single “best guess” number, the method reports a whole RANGE of plausible values, each labeled with how likely it is. You start with an honest “I don't know yet” starting point (called a prior), then you update it using the data you actually collected — more data pulls the range tighter and more confident; less data leaves the range wide and honest about the uncertainty. The specific mathematical shape used for this range is called a Gamma distribution — it's just a flexible curve shape that's good for describing “how many events happened” type numbers, because it can never go below zero (you can't have negative infections) and it can be lopsided (some values are more likely than others).

**WHY THIS CHOICE, NOT SOMETHING SIMPLER?**

Why not just report the single most likely Rt value and leave it there — isn't that simpler and easier to act on? Because a single number invites people to trust it more than it deserves, especially with thin data. A responder who sees “Rt = 3.47” for a 3-case province, with no sense of how shaky that number is, might reasonably panic and divert resources there. A responder who sees “Rt = 3.47, but honestly could be anywhere from 0.1 to 18” makes a very different, better-informed decision. Reporting a range instead of a point is what makes it possible to treat Ituri's tight, trustworthy estimate differently from Sud-Kivu's wide, shaky one — which is exactly what good operational decision-making needs. This is the “be honest about confidence” part of the one goal from before Part 1, showing up for the very first time.

This whole method — echo of past cases, compare to what's happening now, report a range not just one number — is called the Cori method, named after the scientist who developed it. It's used throughout the paper as an independent double-check against the more sophisticated method described next.

**Part 3 · Why One Country-Wide Number Lies to You**

*Matches: Section 2.4, The hierarchical Bayesian renewal model — the heart of the paper*

The Cori method from Part 2 works fine for the whole country, where there's lots of data. But the paper's real question is about SIX separate provinces, and within some of them, dozens of separate health zones. Some of those places have thousands of cases. Others have 3. If you just ran the Cori method separately and independently in every single place, the data-poor places would bounce around wildly — exactly like the 5-coin-flip problem, over and over, everywhere there isn't much data.

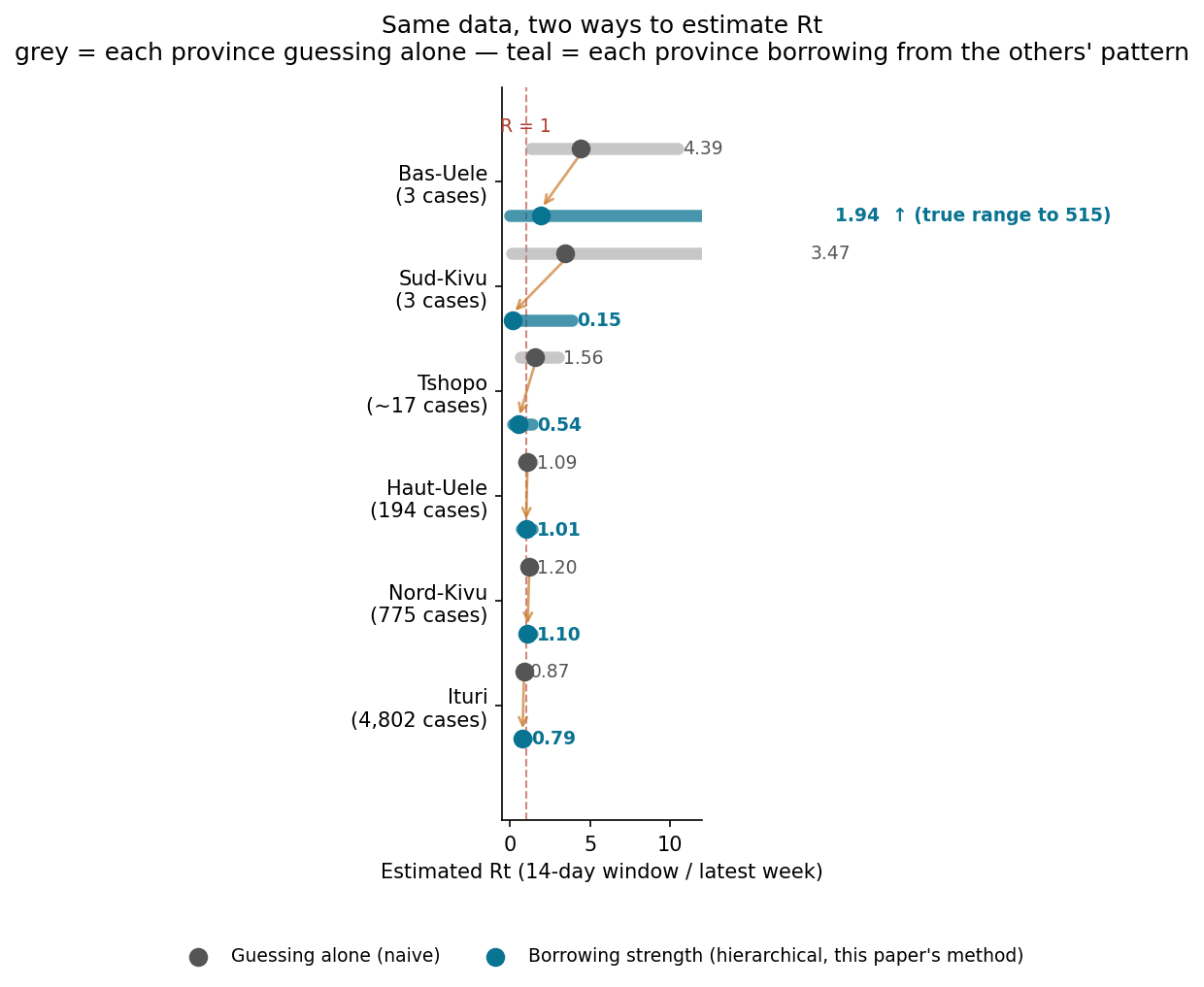

*Figure B — Real numbers from this paper. Guessing alone (grey) vs. borrowing strength (teal), for all six provinces, smallest data at top.*

**WHY THIS CHOICE, NOT SOMETHING SIMPLER?**

This is arguably the single most important “why” in the entire paper, so it's worth spelling out the two ways this could have gone wrong. Option A: force every province to share ONE Rt. That would be simple and stable — and it would make the paper's entire scientific finding literally invisible, since Rwampara declining while Bunia stays elevated would just get averaged away into a single meaningless national number. Option B: estimate every province completely independently, with no connection between them. That would let real differences show up — but it would also let Sud-Kivu's 3 cases produce a wild, meaningless 3.47 estimate that a decision-maker might mistakenly act on. Partial pooling (Part 3's actual method) is the only one of the three options that protects BOTH halves of the goal at once: real differences between places stay visible (Rwampara really does look different from Bunia), while noise from tiny samples gets tamed (Sud-Kivu doesn't get to look falsely alarming). Every other choice described in the rest of Part 3 exists to make this balancing act work.

Look closely at that picture — every number in it is real, taken straight from the paper. Sud-Kivu has only 3 total cases. Guessed alone, its Rt looked like 3.47 (alarmingly high, with an enormous, almost useless range up to 18!). But once it's allowed to “borrow” information from the overall pattern seen across all six provinces, its estimate drops to a far more sensible 0.15. Compare that to Ituri, which has 4,802 cases: guessed alone it's 0.87, and borrowing barely changes it at all, to 0.79. Ituri has enough of its own data that it doesn't need much help from the others.

**The “class photo” analogy**

Imagine you're guessing the average height of kids in your class, but you've only managed to measure 3 kids so far. Do you trust just those 3 measurements completely? Probably not — you'd be smart to also think about “what's a typical height for kids this age, across lots of classes I've seen before,” and let that nudge your guess a bit. As you measure MORE kids in your own class, you'd trust your own class's data more and more, and rely less on the general pattern.

That's exactly what this model does, automatically, place by place. It's called partial pooling — “partial” because it's a middle ground. It doesn't force every province to have the identical Rt (that would hide real, true differences, like the fact that Rwampara and Bunia genuinely are behaving differently right now). But it also doesn't let each tiny province flail around independently with almost no data backing it up. Places with lots of data mostly stand on their own. Places with little data lean more on the overall pattern.

**Rt doesn't stay still — letting it wander, gently, week by week**

Real outbreaks speed up and slow down over time — Rt today isn't necessarily the same as Rt three weeks ago. So instead of assuming one fixed Rt for an entire outbreak, the model lets Rt take a new value every week. But it doesn't let it jump around randomly from nothing each week either — each week's value starts from wherever LAST week's value was, then takes a small, random nudge up or down.

This is called a random walk. Picture a hiker walking down a hill in thick fog: they can't see far, so they take small steps, sometimes drifting left, sometimes right, but they never teleport to a completely different spot on the mountain. Next week's Rt is always close to this week's Rt — that's the assumption, and it matches how real outbreaks actually behave.

**WHY THIS CHOICE, NOT SOMETHING SIMPLER?**

Why not just assume Rt is one fixed number for the whole outbreak, and estimate that single value as precisely as possible? Because that assumption would have been not just imprecise but actively wrong — and it would have hidden the paper's single most important extended finding. Rwampara went from elevated (1.24) to clearly declining (0.53) in three weeks; Butembo went the opposite direction, from declining (0.64) to elevated (1.36). A fixed-Rt model can't represent either of those changes — it would just average them into a meaningless constant. Letting Rt wander week to week is precisely what makes it possible to DETECT that transmission direction itself can reverse, which is a real difference over TIME, the third piece of the one-goal statement from before Part 1.

**A small but important trick: working with the LOG of Rt, not Rt itself**

Rt can never be negative — you can't infect negative-one people. But the small random nudges in a random walk are naturally symmetric (equally likely to go up as down). If you tried to random-walk Rt directly, you could accidentally nudge it below zero, which is meaningless.

The fix: random-walk the LOGARITHM of Rt instead. Without getting into exactly what a logarithm is, here's the part that matters: going from Rt = 2 down to Rt = 1 is “halving” transmission. Going from Rt = 1 down to Rt = 0.5 is ALSO “halving” transmission — same relative change, even though the raw numbers moved by different amounts (2→1 is a drop of 1; 1→0.5 is a drop of only 0.5). On the log scale, both of those “halvings” count as the exact same size of step. That's exactly the property you want for something that naturally changes multiplicatively (by percentages) rather than by fixed amounts.

**WHY THIS CHOICE, NOT SOMETHING SIMPLER?**

This one is a purely technical fix, but it protects something real: without it, the model's “gentle nudge each week” assumption would silently mean something different depending on whether Rt happened to currently be high or low — a nudge of a fixed size is a huge relative change when Rt is near 0.2, but a tiny relative change when Rt is near 5. Working in logs makes “a typical week-to-week change” mean the same thing everywhere on the scale, which is what “gently wander” is supposed to mean in the first place. Get this wrong and the model would implicitly treat small provinces (naturally closer to the edges of plausible Rt) differently from large ones, for no epidemiological reason at all — a hidden, unintended bias, not a real finding.

**Some weeks are just chaotic — accounting for “lumpiness”**

If transmission were perfectly smooth and predictable, you'd expect new case counts to follow a very standard, well-behaved counting pattern (statisticians call this a Poisson distribution). But real outbreaks are lumpy: a single funeral, market day, or treatment-centre cluster can dump in a burst of cases on one particular day, even without the underlying Rt actually changing. A model that doesn't expect any lumpiness will treat every single one of those chaotic days as a five-alarm surprise, even when it's just normal messiness.

The fix is a small extra “how lumpy is this place, generally” knob, called the dispersion parameter (written as the Greek letter phi, φ). It lets the model say: “yes, more cases than expected today — but I already know this place tends to have bursty days, so I won't over-react.”

**WHY THIS CHOICE, NOT SOMETHING SIMPLER?**

Why does “lumpiness” deserve its own knob instead of just letting Rt absorb every bump? Because if you don't model burstiness explicitly, the model has only one way to explain a sudden cluster of cases: decide Rt must have jumped. That would mean single chaotic days — a funeral, a market, a batch of delayed lab results all landing on the same date — could masquerade as genuine changes in transmission. The dispersion knob gives the model a second, more honest explanation (“this place is just naturally bursty”) so that Rt only moves when there's a real, sustained signal behind it. This directly protects “not being fooled by noise” — without it, the paper would risk reporting false alarms every time reporting happened to clump.

**A real decision the authors had to make, and changed their mind about**

Should every province share ONE “how lumpy” knob, or should each province get its OWN? At first, with the smaller dataset, the two choices barely made a difference — so the simpler, one-knob-for-everyone option was used. But once the data grew (more provinces, three more weeks of data, and two zones that dramatically reversed direction), the evidence shifted: separate knobs for each province genuinely did a better job predicting held-out data (Part 5 explains exactly how “better” gets measured). So the authors switched to separate knobs — and said so plainly in the paper, rather than quietly keeping the old choice. That's what good science is supposed to look like: the answer is allowed to change when better evidence shows up, as long as you're honest about it changing.

**WHY THIS CHOICE, NOT SOMETHING SIMPLER?**

Why bother letting the lumpiness knob vary by province at all, instead of just always using one shared value for simplicity? Because a bigger city with steady reporting infrastructure and a remote zone with irregular reporting genuinely don't have the same amount of natural burstiness — forcing them to share one value means the model's uncertainty ranges are wrong for BOTH: too narrow somewhere, too wide somewhere else. This is really the same three-part goal again, at a more technical level: letting each place have its own dispersion is what makes the reported uncertainty honest FOR THAT SPECIFIC PLACE, not just honest on average across all of them.

**Part 4 · How Does a Computer Actually FIND These Numbers?**

*Matches: Section 2.4 and 2.10, Estimation and software*

Here's a problem: the model in Part 3 has HUNDREDS of unknown numbers tangled together — a separate Rt for every province, for every week, plus the lumpiness knobs, plus how much each province leans on the group pattern. There's no simple formula you can just plug numbers into and get an answer, the way you can with y = 2x + 3. It's too tangled for that.

**The extremely smart guessing game**

Instead, the computer plays a guessing game called Markov Chain Monte Carlo (don't worry about the name). Here's roughly what it does:

**1.** Start with a random guess for every unknown number.

**2.** Nudge the guess slightly in a direction that fits the real data a bit better.

**3.** Occasionally, on purpose, take a step that looks slightly WORSE — this stops the computer from getting stuck confidently believing one narrow, possibly-wrong answer.

**4.** Repeat this millions of times.

**5.** Look at everywhere the guessing game visited. The places it visited most OFTEN are the most believable answers.

A good way to picture this: imagine a marble rolling around inside a bowl-shaped valley, with a bit of random jiggling. It won't sit perfectly still at the exact bottom, but it will spend most of its time near the bottom, and only rarely wander up near the rim. If you tracked everywhere that marble went and made a map of “how often was it here,” the map would look like a hill centered on the true lowest point — and that's basically the shape of an answer this method gives you: not one number, but a whole hill of plausible numbers, tallest where the model is most confident.

The specific, clever version of this guessing game used in the paper is called the No-U-Turn Sampler (NUTS) — a smarter version that's good at not wasting time wandering back over places it's already checked.

**WHY THIS CHOICE, NOT SOMETHING SIMPLER?**

Why go to all this trouble instead of using a faster, simpler estimation method? Because every simpler method (the kind that gives you one formula and one answer) requires assuming the tangled web of unknowns can be pulled apart cleanly — and in this model, they genuinely can't be: each province's Rt trajectory partly depends on the shared pooling pattern, which itself depends on every OTHER province's data. MCMC is slower, but it's honest about the tangle instead of assuming it away. Skipping this step and using a simpler shortcut would mean quietly getting either the point estimates or — more likely — the uncertainty ranges wrong, without any obvious sign that anything had gone wrong.

**Checking the computer didn't get stuck somewhere weird**

Since this is a random guessing process, how do you know it actually worked? The trick: run the ENTIRE guessing game several times over, completely independently, starting from different random spots (these independent runs are called chains — the paper uses 2 or 4 of them depending on the model). If all the chains end up wandering around the same neighborhood, that's a great sign they all found the same real answer. If one chain got stuck somewhere completely different, that's a red flag.

The number used to check this is called r-hat. Without the formula: r-hat close to 1.00 means all the independent chains agree with each other. In this paper, most models had r-hat right at 1.00; the busiest zone-level models (with up to 25 separate places being estimated at once, and fewer chains run to save computing time) had r-hat up to about 1.02 — still good, but the paper says so honestly rather than hiding it, since it's slightly above the very strictest bar the original, smaller analysis achieved.

**WHY THIS CHOICE, NOT SOMETHING SIMPLER?**

Why not just trust whatever the computer spits out after running once? Because a guessing game that hasn't been checked could easily be reporting an answer that reflects where it happened to get stuck, not the true shape of the evidence — and a single run has no way to tell you which one happened. Running multiple independent chains and checking they agree is the only way to catch this kind of silent failure. This protects the credibility of literally every number in the paper: r-hat isn't a result anyone cares about for its own sake, but without checking it, none of the OTHER results could be trusted either.

**Part 5 · How Do We Know We Can Actually Trust It?**

*Matches: Section 2.5–2.6 and 3.3, Model validation*

Getting an answer out of a computer doesn't automatically make it a good answer. The paper runs three separate checks — think of them as three different kinds of exam the model has to pass.

**Check 1: the flashcard test (leave-one-out cross-validation)**

Imagine you're studying for a vocabulary test with 100 flashcards. A good way to check if you've actually LEARNED the pattern (not just memorized the specific cards) is: hide one flashcard, see if you can still guess it correctly using everything else you know, then put it back and try a different hidden card. Repeat for every card.

The model does exactly this with real data points: hide one, ask the model (trained on everything else) to predict it, and measure how surprised the model is by the real answer. Do this for every data point, then average the surprise. This lets you compare two versions of the model fairly — for example, “one lumpiness knob for everyone” versus “one lumpiness knob per province.” Whichever version is LESS surprised, on average, when tested this way, is genuinely doing a better job — not just fitting the data it already saw, but actually capturing the real pattern.

The paper reports this “average surprise difference” as Δelpd (don't worry about the name — bigger means a bigger difference). At the original cutoff, the difference between the two lumpiness-knob options was tiny (0.3, with a typical wobble size of about 1.7) — too small to trust as real, so the simpler option was used. At the extended cutoff, the difference grew to 5.51 (wobble size about 3.44) — now clearly bigger than the normal wobble, so the more detailed option was adopted instead.

**WHY THIS CHOICE, NOT SOMETHING SIMPLER?**

Why not just always use the more detailed, more flexible version of the model — extra flexibility can only help, right? Actually, no: a model with more knobs to turn can fit the SPECIFIC data it already saw better almost by definition, even if the extra flexibility is just chasing noise rather than capturing anything real (this is called overfitting). The flashcard test is the safeguard against fooling yourself this way — it only rewards the more complex model if that complexity genuinely helps predict data it HASN'T seen yet. This protects the goal's “not being fooled by noise” clause in a very specific way: it stops the researchers themselves from being fooled by their own model looking impressively good on data it was trained on.

**Check 2: does the model's own “pretend data” look like the real data?**

Once the model has learned its rules, you can ask it to pretend to generate brand-new, fake outbreak data using only those learned rules — no peeking at the real numbers. If the model is good, the REAL data should usually fall comfortably within the range of “pretend” data it generates. Specifically, since the paper asks for a 95% range, the real data SHOULD land inside that range about 95% of the time — not 60%, not 100%.

This is called a posterior predictive check, and the measurement is called coverage. The model in this paper achieved 96.1% coverage — very close to the 95% target, meaning the ranges it reports are honestly sized, not too narrow (overconfident) and not too wide (uselessly vague).

A helpful comparison: a weather forecaster who says “70% chance of rain” is well-calibrated if, out of all the times they say that, it actually rains about 70% of the time — not 20% and not 100%. Coverage checks the exact same kind of honesty, but for this Rt model.

**WHY THIS CHOICE, NOT SOMETHING SIMPLER?**

Why isn't it enough for the model's BEST-GUESS numbers (the medians) to look reasonable — why specifically test the RANGES too? Because a model can get the middle of the range roughly right while still being badly miscalibrated about its own uncertainty — too confident (ranges too narrow, so real data keeps surprising it) or too timid (ranges too wide, so every result looks equally uncertain and useless for decisions). Since almost every number in this paper is reported as a range, not a bare point estimate, this check is really testing the part of the output that readers will actually use to make decisions — directly protecting the “be honest about confidence” clause of the one goal.

**Check 3: what if a key assumption was wrong?**

Remember from Part 2 that the model assumes a typical 15.3-day delay between one infection and the next (the generation interval) — but nobody has measured this exactly for THIS specific outbreak. It's a reasonable estimate borrowed from similar past outbreaks, but it's still a guess.

So the authors asked: what if that guess is wrong? They reran the ENTIRE model twice more — once assuming a shorter typical delay, once assuming a longer one — and checked whether the province RANKING changed (which province looks worst, second-worst, and so on), even if the exact numbers shifted a little.

The tool for comparing rankings (rather than exact values) is called Spearman rank correlation — it asks “did the order stay the same,” not “did the exact numbers stay the same.” A value of 1.0 means the order was IDENTICAL every time, no matter which delay assumption was used. That's what the paper found, both times, which means the main conclusions aren't secretly hanging on a shaky guess about the delay.

**WHY THIS CHOICE, NOT SOMETHING SIMPLER?**

Why test RANKING instead of demanding the exact Rt numbers stay identical under a different delay assumption? Because demanding exact stability would be an unreasonably strict, and slightly beside-the-point, standard — changing an input assumption SHOULD shift the precise numbers somewhat; that's expected and fine. What actually matters for the paper's real-world use (deciding where to focus response resources) is whether the RELATIVE priority between places changes. Testing rank rather than exact value is choosing the test that matches how the result will actually be used — a general lesson worth remembering: always check robustness in the units that matter for the decision, not just in whichever units are easiest to test.

**Part 6 · An Experiment, Not Just an Argument**

*Matches: Section 2.9 and 3.5, Generalisability validation*

Back in Part 1, we saw that “connect the dots” (interpolation) is a better fix for reporting gaps than “freeze the score, then dump it all at once” (forward-fill). But that fix was discovered by looking at ONE specific gap, in this ONE specific outbreak. How do we know it isn't just a coincidence that worked out for this particular case?

The authors ran a real experiment to check. Here's exactly what they did, step by step — no advanced statistics needed to follow this part:

**1.** They found a completely different, older outbreak (2018–2020, a different part of the Congo) where the true, complete data is already known — no mystery, no missing days.

**2.** They picked one well-measured place from that old outbreak (a town called Beni) during a period when cases were clearly rising.

**3.** They deliberately DELETED two weeks of that clean data, pretending they didn't know it — on purpose creating exactly the same kind of gap that caused trouble in the real BDBV data.

**4.** They rebuilt the missing two weeks TWO different ways: freeze-and-dump (forward-fill) and connect-the-dots (interpolation).

**5.** Because they still secretly had the TRUE numbers (from step 1), they could compare each rebuilt guess against the real answer and measure exactly how wrong each method was.

**The result**

Freeze-and-dump was off by a total of 110 cases across the affected weeks. Connect-the-dots was off by only 21 — about five times better. And because this test used a totally different outbreak, town, and time period from anything else in the paper, this isn't just “it worked once, by luck.” It's real evidence that the fix generally works whenever this particular kind of reporting gap happens.

**WHY THIS CHOICE, NOT SOMETHING SIMPLER?**

Why wasn't it enough to just explain the interpolation fix clearly and let readers accept the reasoning, the way Part 1 originally did? Because a plausible-SOUNDING fix and a fix that's actually TESTED against ground truth are different levels of evidence, and good science tries to tell them apart rather than assuming a good argument is the same as a good result. This experiment is the one piece of the paper that doesn't rely on trusting the big hierarchical model at all — it's independent, simple, and checkable by anyone with the same two datasets. That independence is exactly what makes it strong evidence rather than just a good story: it could have come out the other way (forward-fill winning, or the two methods tying), and the fact that the authors ran it anyway, willing to be wrong, is itself part of what makes the result trustworthy.

This is genuinely one of the simplest ideas in the whole paper, and also one of the most convincing, precisely because it doesn't rely on trusting a complicated model — it's just: break something on purpose where you already know the right answer, try your fix, and measure how close you got.

**Part 7 · What Does “95% Credible Interval” Actually Mean?**

*Used constantly throughout the paper — every single Rt number comes with one*

You'll see something like this everywhere in the paper: “Rt = 0.79 (95% CrI 0.61–1.04).” The middle number (0.79) is the single best guess. The two numbers in brackets are the credible interval — the range the model thinks is very likely to contain the real answer, based on everything it was told (the data, the generation-interval assumption, all of it).

A useful, honest way to think about it: “Given everything we assumed and everything we measured, we think there's about a 95% chance the true Rt is somewhere between 0.61 and 1.04.” The WIDTH of that range tells you how confident to be. A narrow range means the model has a lot of good data to work with. A wide range is the model being honest about not knowing very much yet.

**Why the same country has both very narrow and very wide ranges**

Look back at Figure B in Part 3. Ituri's range (0.61–1.04) is fairly tight, because it has 4,802 cases behind it — lots of evidence. Bas-Uele's true range stretches all the way past 500 (shown capped in the figure for readability) — because it only has 3 cases so far, so the model is mostly just saying “I genuinely don't know yet, and I'm not going to pretend otherwise.”

That's actually a feature, not a flaw. A model that confidently reported a narrow range for a 3-case province would be lying to you. Being appropriately uncertain, out loud, when the data doesn't support confidence, is exactly what a trustworthy method should do.

**WHY THIS CHOICE, NOT SOMETHING SIMPLER?**

Why does a paper about disease transmission spend so much effort on ranges, when a single decisive number would be so much easier for a response coordinator to act on? Because reporting a false sense of precision is a real operational hazard, not just a statistical nicety: if Bas-Uele's true range (0.01 to over 500) got collapsed into a single confident-looking “1.94,” a reader could easily mistake early, thin data for a settled answer and misallocate scarce response resources on that basis. An honest wide range is a signal in its own right — it says “watch this place, but don't over-read it yet” — which is genuinely more useful for decision-making than false confidence, even though it feels less satisfying to read. This is the “be honest about confidence” clause of the one goal, showing up for the last time, now fully connected to why it mattered from the very first page.

**Part 8 · If You Only Remember Five Things**

**1.** Rt above 1 means an outbreak is growing; below 1, it's shrinking. Every part of this paper exists to pin down that one number, more precisely, in more places, more recently.

**2.** A single national average can hide the fact that some places are getting better while others are getting worse — or that the SAME place can flip from one to the other within a few weeks.

**3.** Places with little data shouldn't be trusted to “guess alone” — they do better “borrowing strength” from the overall pattern seen everywhere else, while places with lots of data can mostly stand on their own.

**4.** Every number in this paper comes with an honest range, not a fake-precise single value — and the range gets appropriately wider wherever there's genuinely less to go on.

**5.** The authors didn't just claim their methods and fixes work — they tested them: against held-out data, against pretend simulated data, against alternate assumptions, and against a completely different outbreak. Every one of those tests is a chance for the method to fail publicly, and the paper reports what happened either way.

**One last thing**

If anything in this document made a piece of the real paper click that didn't before, that's the entire point. If something is STILL confusing after reading this, that's not you being bad at math — it almost certainly means I explained it badly, and it's worth asking about directly rather than assuming it's over your head. Feynman's whole point was that if an explanation doesn't work, the fault is in the explanation, not the listener.
